# SAFE REOPENING STRATEGIES FOR EDUCATIONAL INSTITUTIONS DURING COVID-19: A DATA-DRIVEN AGENT-BASED APPROACH

**DOI:** 10.1101/2020.09.04.20188680

**Authors:** Ujjal K mukherjee, Subhonmesh Bose, Anton Ivanov, Sebastian Souyris, Sridhar Seshadri, Padmavati Seshadri, Ronald Watkins, Yuqian Xu

## Abstract

Many educational institutions have partially or fully closed all operations to cope with the challenges of the ongoing COVID-19 pandemic. In this paper, we explore strategies that such institutions can adopt to conduct safe reopening and resume operations during the pandemic. The research is motivated by the University of Illinois at Urbana-Champaign’s (UIUC’s) SHIELD program, (https://www.uillinois.edu/shield), which is a set of policies and strategies, including rapid saliva-based COVID-19 screening, for ensuring safety of students, faculty and staff to conduct in-person operations, at least partially. Specifically, we study how rapid bulk testing, contact tracing and preventative measures such as mask wearing, sanitization, and enforcement of social distancing can allow institutions to manage the epidemic spread.

**Research Design:** This work combines the power of analytical epidemic modeling, data analysis and agent-based simulations to derive policy insights. We develop an analytical model that takes into account the asymptomatic transmission of COVID-19, the effect of isolation via testing (both in bulk and through contact tracing) and the rate of contacts among people within and outside the institution. Next, we use data from the UIUC SHIELD program and 85 other universities to estimate parameters that describe the analytical model. Using the estimated parameters, we finally conduct agent-based simulations with various model parameters to evaluate testing and reopening strategies.

**Results:** The parameter estimates from UIUC and other universities show similar trends. For example, infection rates at various institutions grow rapidly in certain months and this growth correlates positively with infection rates in counties where the universities are located. Infection rates are also shown to be negatively correlated with testing rates at the institutions. Through agent-based simulations, we demonstrate that the key to designing an effective reopening strategy is a combination of rapid bulk testing and effective preventative measures such as mask wearing and social distancing. Multiple other factors help to reduce infection load, such as efficient contact tracing, reduced delay between testing and result revelation, tests with less false negatives and targeted testing of high-risk class among others.

**Contributions:** This paper contributes to the nascent literature on combating the COVID-19 pandemic and is especially relevant for educational institutions and similarly large organizations. We contribute by providing an analytical model that can be used to estimate key parameters from data, which in turn can be used to simulate the effect of different strategies for reopening. We quantify the relative effect of different strategies such as bulk testing, contact tracing, reduced infectivity and contact rates in the context of educational institutions. Specifically, we show that for the estimated average base infectivity of 0.025 (*R*_0_ = 1.82), a daily number of tests to population ratio *T/N* of 0.2, i.e., once a week testing for all individuals, is a good indicative threshold. However, this test to population ratio is sensitive to external infectivities, internal and external mobilities, delay in getting results after testing, and measures related to mask wearing and sanitization, which affect the base infectivity.

## 1. Introduction

The majority of the educational institutions in the United States, ranging from primary schools to universities, have temporarily ceased in-person classes and other activities due to the ongoing COVID-19 pandemic. While the importance of reopening is widely recognized,^1^ there is lack of consensus on the strategies necessary to safely reopen these institutions.^2^ The Center for Disease Control (CDC) has issued reopening guidelines that include extensive hand hygiene, cloth face coverings, repetitive disinfection, physical barriers and spacing of individuals inside enclosed surroundings, frequent testing, etc.^3, 4^ Sharp increase in COVID-positive cases from reopening with in-person interactions prompted eventual re-closures.^5^ For example, Cherokee County School District in the state of Georgia, USA, quarantined 250 staff members and students after reopening in August, 2020.^6^ Similarly, the University of North Carolina, Chapel Hill, USA, canceled in-person classes after finding >130 confirmed infected cases in the very first week after reopening.^7, 8^ Motivated by these observations, we explore the question of whether educational institutions and other organizations can safely commence in-person operations amidst the COVID-19 pandemic. In particular, we identify measures that are necessary to ensure the safety of the members of an institution and the public at large. To do so, we employ a combination of analytical modeling, data analysis and agent-based simulation. We first develop a mathematical model that captures the dynamics of the infection process with bulk testing and contact tracing. Then, we estimate some of the analytical model parameters from real data from a number of universities in the United States. Finally, we use the parameter estimates to conduct an agent-based simulation experiment to evaluate strategies for safe reopening.

SARS-CoV-2 is a novel strain of coronavirus that currently does not have an approved cure.^9–11^ For mitigation, a variety of strategies have been implemented across the globe, ranging from complete lock-down of large geographical areas^12^ to partial restrictions on mobility and mask enforcement in public places.^13^ A particular challenge associated with this virus is its asymptomatic transmission in which many infected individuals remain asymptomatic from a few days to several weeks and yet transmit the disease to susceptible people.^14, 15^ We mention the results from Hao et al. (2020)^16^ to highlight the seriousness of asymptomatic transmission.^17^ As Ceylan (2020)^18^ reveals, Italy’s infected population may have ranged between 2.2 and 3.5 million in number as of May 4, 2020, while detected infections numbered a mere 200K. The potency of asymptomatic transmission is no different within an educational institution. Thus, we posit that a reopening strategy is difficult to design without the ability to conduct rapid bulk testing (testing everyone once every few days) so that one can detect and arrest the spread of infections through systematic isolation and quarantining of those who test positive for infection. Our work is motivated and guided by the SHIELD program of the University of Illinois at Urbana-Champaign (UIUC). In this program, the university is currently testing >10K students and staff every day (that amounts to 0.2 tests per individual per week) through saliva-based tests.

There are multiple testing options for COVID-19. The nasal swab-based tests that utilize reverse-transcriptase polymerase chain reaction (RT-qPCR) define the gold standard for testing of COVID-19 and is acknowledged to be very accurate.^19, 20^ The US Food and Drug Administration (FDA) has recently approved saliva-based rapid testing^21^ that utilizes a loop-mediated isothermal amplification (LAMP) technique (see Wyllie et al. (2020)^22, 23^). LAMP tests are significantly less costly than RT-qPCR tests, as Augustine et al. (2020)^24^ reveals. In the case of the UIUC SHIELD program, results from the LAMP tests are being made available within 6 to 12 hours for upwards of 10K daily tests. A recent study in the New England Journal of Medicine^22^ compared the sensitivities of the RT-qPCR tests with the saliva-based LAMP tests on 70 confirmed COVID-19 patients. They found that LAMP tests were able to detect a higher number of SARS-CoV-2 RNA samples (5.58 with 95% CI of 5.09 to 6.07) as compared to the nasal swab-based RT-qPCR tests (4.93 with 95% CI of 4.53 to 5.33) within 1 to 3 days of the infection. Moreover, a higher percentage of COVID-19 patients tested positive up to 10 days in saliva-based LAMP testing. The study also found that LAMP tests exhibited equal or higher sensitivity compared to RT-qPCR tests when patients are asymptomatic. While both RT-qPCR and LAMP testing procedures require similar laboratory turnaround time, sample collection and testing for saliva-based testing is faster and can be more easily administered en masse. The cost of the SHIELD saliva based testing is between $20 and $30 (https://www.uillinois.edu/shield) compared to more than $100 per nasal swab test. For bulk testing at large universities and institutions cost considerations are equally important as accuracy of testing.^25^ In this paper, we consider two different channels of testing for an educational institution–one conducted in bulk at regular intervals and another through tracing of contacts of individuals who already test positive. While our results will continue to hold for other testing options, our study is motivated by the saliva-based testing paradigm adopted by UIUC.

Methodologically, we develop an analytical epidemic model with testing, use data to conduct parameter estimation and run agent-based simulation experiments to evaluate viable reopening strategies for educational institutions. Compartmental diffusion models have been widely employed to study epidemic processes, dating back to Serfling (1952).^26^ However, without modification, such models are not suitable for the analysis of COVID-19 infections among small populations such as those in educational institutions.^27, 28^ Therefore, we propose our own compartmental model that accounts for the asymptomatic transmission of COVID-19, reflects the impact of testing on infection transmission, considers the effect of small populations as that of educational institutions and incorporates the role of infections in the counties surrounding the institution. The model has several parameters that describes the infection and testing process. We use a nonlinear regression technique that estimates these parameters from data. For estimation, we use data from COVID-19 dashboards maintained by several educational institutions within the US, including that from the UIUC SHIELD program. Using these estimated parameters, we devise an agent-based simulation experiment.^29^ This experiment utilizes simple probabilistic rules for contact, infection transmission, testing and recovery, that taken together, seeks to mimic reality and build a digital twin for the epidemic process in practice. Testing and reopening strategies are then evaluated against the random sample path outputs of the agent-based simulation. This experimental setup makes our policy recommendations quite robust to parameter variations and assumptions made to derive the compartmental model.

Our results shed light on viable strategies that institutions can adopt to cope with the challenges of the epidemic and yet, prioritize the health and safety of their members. First, we argue that since COVID-19 is characterized by asymptomatic transmission, rapid bulk testing is vital to safe reopening of educational institutions. However, without proper mask enforcement and social distancing will require testing almost every individual every day. The size of an educational institution makes it imperative that they conduct bulk testing and enforce precautionary measures in tandem to effectively manage testing costs to resume in-person activities. Second, we provide a framework to analyze the allocation of testing capacity between bulk testing and contact tracing. We demonstrate that the value of contact tracing is, somewhat counter-intuitively, higher when the positivity rate from bulk testing is low. With low positivity rates (e.g., during the initial stages of the epidemic), the probability of discovering infected individuals from bulk testing remains low, when contact tracing provides a targeted mechanism to discover infected individuals. As the infection spreads, development of bulk testing capabilities becomes crucial for effective mitigation of the infection spread. Instead of adopting a fixed allocation between bulk testing and contact tracing, a flexible and adaptive allocation based on estimated positivity rates of testing is shown to be more cost-efficient. We show that an institution must test more during the initial stages of reopening. The testing levels can then be ramped down adaptively as the infection load (positivity rate) decays. At UIUC, upon reopening in August 2020, students were required to test twice a week and faculty and staff were required to test once a week, however, after a few weeks students were moved to three times a week and others were moved to twice a week testing due to increasing infectivity within UIUC. Once the infections dampened by the middle of September, the frequency of testing for students and faculty was moved back to twice and once per week respectively. Third, we show that fast revelation of testing results along with measures to isolate detected positive individuals plays a rather central role in designing reopening strategies. In other words, it is important to quickly identify infected individuals and restrict them from further spreading the disease among the susceptible population. The rapid saliva-based LAMP tests with an average turnaround time of 6-12 hours (e.g., the testing mechanism of the UIUC SHIELD program) are ideally suited for this task. The inability to do so (particularly when the delay grows beyond a day) renders testing largely ineffective. Fourth, we demonstrate that testing different sub-populations (based on risk categories) can be an important policy consideration. This finding supports the efforts of several universities such as UIUC that are testing the student population and the faculty/staff population at different frequencies. Students on campus at UIUC are being tested twice every week and faculty/staff are tested once every week for building and facility access, and this frequency of testing was changed over time to adapt to changing internal and external infections. Fifth, our data analysis reveals that the higher the infection rate of the county where a university is located, the higher is the infection rate within the university. The relationship is in fact dyadic in that large universities with a significant influx of students from outside, contribute significantly towards the growth of infection in the surrounding region. Thus, considering the infection spread within an educational institution in isolation cannot reveal the whole story; the prevalence of the disease in its vicinity plays an important role in this dynamics.

The rest of the paper is organized as follows. In §2, we describe the methods including the analytical model that describes the infection process, testing and contact tracing; the data and the parameter estimation process, and the simulation setup. In §3, we present and discuss the results from our empirical analysis. Finally in §4, we conclude the paper with remarks on our results and directions for future work.

## 2. Methods

We begin by describing an epidemic model that accounts for the infection process as well as the testing process. Then, we describe the data collected from several US universities and explain how we estimate the model parameters from this data. Finally, we use the parameter estimates and the analytical setup to conduct an agent-based simulation to elicit feasible strategies for reopening. Sensitivities of our strategies to variations in the key assumptions of our model are also described.

### 2.1. The Epidemic Model

We consider an educational institution with a population (total number of members) *n*. To model the epidemic dynamics, we segment the population into different classes and track the dynamics of the number of members in each class. Specifically, we consider the following population segments on day *t*:

- Susceptible individuals, *s*_*t*_, who are not infected but can get infected when they come in contact with infected individuals.
- Infected but undetected individuals, *u*_*t*_, who can infect susceptible individuals when they come in contact with them.
- Infected individuals who test positive, *p*_*t*_.
- Individuals who became COVID-19 positive but ultimately recovered from it, *r*_*t*_.

To streamline the modeling process, we consider a specific sequence of events on day *t*. Many of the events we describe occur simultaneously. The sequence, however, helps us to formally describe the process quite easily without losing the essential features of the epidemic process. Specifically, consider the following event sequence: (i) The pool of all contacts (*c*_*t*−1_ in number) who came in contact with COVID-19 positive individuals on day *t* − 1 are tested and then a portion of the institution population is tested under the institution’s bulk testing policy, (ii) members of the institution, as a result of their daily affairs, come in contact with other individuals within and outside the institution that results in new infection transmission, (iii) test results arrive, all COVID-19 positive individuals are isolated, and a list of contacts of all COVID-19 positive individuals is created to be tested the following day, and (iv) some infected individuals recover. In what follows, we develop a mathematical model to describe the dynamics of the population segments through this sequence of events:

#### Step 1. Testing traced contacts from day *t* − 1 and bulk-testing

Let, *T*_*t*_ denote the testing capacity on day *t* that is assumed to exceed *c*_*t*−1_, the number of contact traced individuals. First, all *c*_*t*−1_ individuals are tested. Then, the balance of *T*_*t*_ − *c*_*t*−1_ tests remaining after testing the contacts of detected infections, are used for bulk testing. Let, testing *c*_*t*−1_ individuals through contact tracing result in 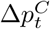 new positive cases, and bulk testing *T*_*t*_ − *c*_*t*−1_ people result in 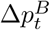 new cases. We now derive an expression for 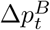 and leave un-derived the expression for 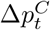 till we have described step 3. Following the protocols of the UIUC SHIELD program, we assume that bulk testing is conducted at a predetermined regular frequency among the mobile part of the population that comprises susceptible, undetected infectious and recovered individuals. Detected positive individuals are assumed to be isolated. The size of this mobile population is *n*_*t*_ := *s*_*t*_ + *u*_*t*_ + *r*_*t*_ = *n* − *p*_*t*_. We assume that positive test results from bulk tests on day *t* within the tested population occurs at the same frequency as the ratio of undetected positive individuals to the total susceptible population among the mobile part of the population. Such a premise is justified when the institution and the testing capabilities within the institution are large enough. For the UIUC SHIELD program, we expect this assumption to be valid, given that UIUC has 50K members and are conducting > 10*K* daily tests. Thus, the (expected) number of new infections detected through bulk testing is given by

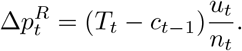

Notice that test results in our model do not arrive before the population segments interact in day *t* and give rise to new infections. As a result, the number who test positive on day *t* depends only on the size of the population segments at the start of day *t*.

#### Step 2. Infection propagation through contacts

Susceptible, undetected infected and recovered individuals interact. These interactions result in new infections. At time *t*, let 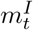 denote the number of members that each individual within the institution meets within the institution. Thus, 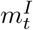 is a measure of intra-institution mobility or contact rate. Similarly, we encode the contact rate between members of the institution and the public at large in the vicinity of the institution in 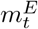. The contact rates depend largely on the nature and the frequency of activities that include in-person classes, office meetings, commercial activities, etc. The interactions create opportunities of infection transmission both from internal and external sources. Let new infections get transmitted at a rate 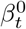 when a susceptible person within the institution meets an infected individual either within the institution or outside of it at time *t*.^1^ In addition to the inherent nature of the virus, this rate depends on the extent to which people adopt preventative measures such as mask wearing and social distancing, that may vary over time. We now count the expected number of new infections that result from interactions within the institution and outside of it.

##### Infection growth from intra-institution contacts

Consider a susceptible individual at time *t* that meets 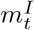 people within the population at time *t*. The probability that *k* among them are infected is given by 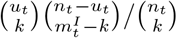 and the probability that at least one among them infects this individual upon interaction is 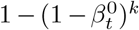. Here, the notation 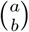 denotes the number of ways of choosing *b* objects from a collection of *a* objects without replacement. Multiplying the above probabilities and summing over *k* yields the probability of a new infection to be 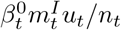, when *β*^0^ is assumed small (*β*^0^ ≈ 1 − 8% as estimated from data from US universities in §2.2). See Appendix A.2 for a detailed derivation. The average growth in the number of infections within the institution due to interactions within the institution therefore becomes 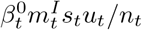.

##### Infection growth from external contacts

A very similar calculation for contacts outside of the institution leads to 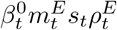 new infections. Here, 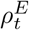 is the positivity rate of COVID-19 infections among people outside of the institution but in the vicinity of the institution. See the Appendix A.2 for the derivation. This rate plays a similar role in the expression for infection growth from external contacts as *u*_*t*_*/n*_*t*_ plays in the expression for intra-institution contacts.

We approximate the total number of new infections by the sum of infections from within and outside of the institution, given by 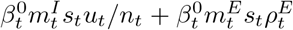. This assumption is justified, and the expression in formally derived in the Appendix A.2. As a result of these contacts, the above number of individuals migrate from the susceptible segment to the undetected infected group.

#### Step 3. COVID-positives are isolated, contact pool is created

Upon receiving the test results, individuals who test positive are isolated. As a result, they no longer contribute to further infection-spread. The number of COVID-positive patients increases by Δ*p*_*t*_. Recall that bulk testing *T*_*t*_ − *c*_*t*−1_ individuals results in 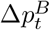 newly detected cases, for which we already derived an expression. Contact tracing *c*_*t*−1_ individuals results in 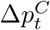 new positive cases, for which we now derive an expression. Let *P* denote the probability that a person in the contact pool of detected positive individuals on day *t* − 1 is infectious by the end of day *t* − 1. Then, the expected number of new cases detected through contact tracing will be 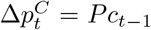. To estimate *P*, notice that an individual in the contact pool was either already infected at the start of day *t* − 1, or got newly infected through interactions on day *t* − 1. The probability that an individual in the contact list started day *t* − 1 being in the undetected infected group should roughly equal the fraction of that group within the mobile population within the institution, given by *u*_*t*−1_*/n*_*t*−1_. The probability of finding an individual who got infected on day *t* − 1 is 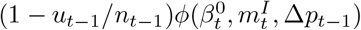. The exact expression for *ϕ* is included in Appendix A.3, where we show that it is reasonable to approximate it by 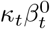, where

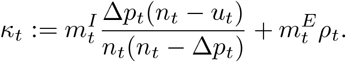

This approximation is valid when Δ*p*_*t*−1_ « *u*_*t*−1_ « *n*_*t*−1_. We expect that regime to hold in practice from the UIUC SHIELD program, given that Δ*p* < 100 and *n* ≈ 50*K*. We do not directly observe the sequence of *u*_*t*_’s; however, we expect the number to be more than 5Δ*p*, given that every individual tests at least once per week. Upon collecting the terms for *P*, we get

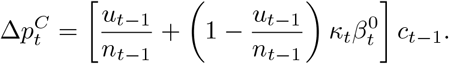

Thus, the total number of detected infections on day *t* becomes 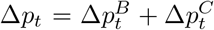. Next, we estimate the expected number of contacts *c*_*t*−1_ of those who were detected as infected on day *t*−1.

Pick a mobile individual within the institution that is not part of the group that newly tested positive on day *t* − 1. The total number of such individuals is *n*_*t*−1_ −Δ*p*_*t*−1_. Then, *c*_*t*−1_ equals this number multiplied by the probability that they belong to the contacts of the newly COVID-positive group Δ*p*_*t*−1_. Contact tracing is either conducted manually (e.g., through interviews, phone calls, etc.) or through automated means (e.g., through a cell phone application such as the Safer Illinois app employed by UIUC). Perfectly tracing all contacts of a positive individual is challenging. The accuracy of tracing depends on several factors, such as the patient’s recollection of contacts, record-keeping at locations where the patient may have visited, adoption and usage of mobile apps, etc. Let 0 < *η* < 1 model the efficiency of the contact tracing process.

Then, the expected size of the contact pool, per our description, becomes

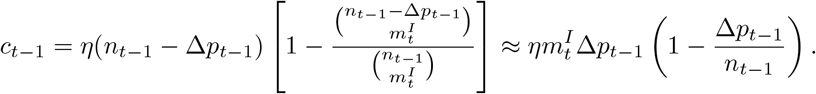

The approximation is derived in the Appendix A.3 under the assumption that 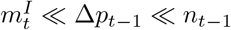. Again, we expect this inequality chain to be valid for UIUC, given that measures related to social-distancing and mask wearing are relatively strictly enforced in an institutional setup. In the case of the UIUC and other similar institutions, usually, 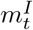 is below 10, Δ*p*_*t*_ ranges between a few tens to a few 100’s, and *n*_*t*_ is in thousands, as we will see in greater detail in section 3.

#### Step 4. Some infected individuals recover

The average time to recover from COVID-19 has been computed to be between 12 and 15 days after which the infected individuals move to the recovered group. The average recovery rate *γ* is assumed to be the inverse of the average time to recover. Incidence of repeat infections are rare. Hence, we deem the recovered group as no longer infective or susceptible to infection. For an educational institution with bulk testing capabilities such as UIUC, we only model the recovery process for individuals who test positive at some point. That is, we do not allow undetected infected population to recover. Such an assumption is justified in the presence of bulk testing such as the SHIELD program of UIUC that tests all individuals at least once a week, thus, identifies all undetected cases. The recovery process is mathematically modeled in

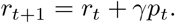

#### 2.1.1. Summary Of The Epidemic Model

The infection dynamics due to transmission, recovery and testing as illustrated in Figure 1 results in the following updates to the population segments within the institution. In Figure 1, we show the four distinct stages that an individual can be in at any time, i.e., susceptible, undetected infected, positive, and recovered, the dynamics of which is mathematically captured in 2.1. Additionally, some individuals from the susceptible and the undetected infected group may be indicated for contact tracing, who either move to positives or susceptibles after getting tested.

**Figure 1.**
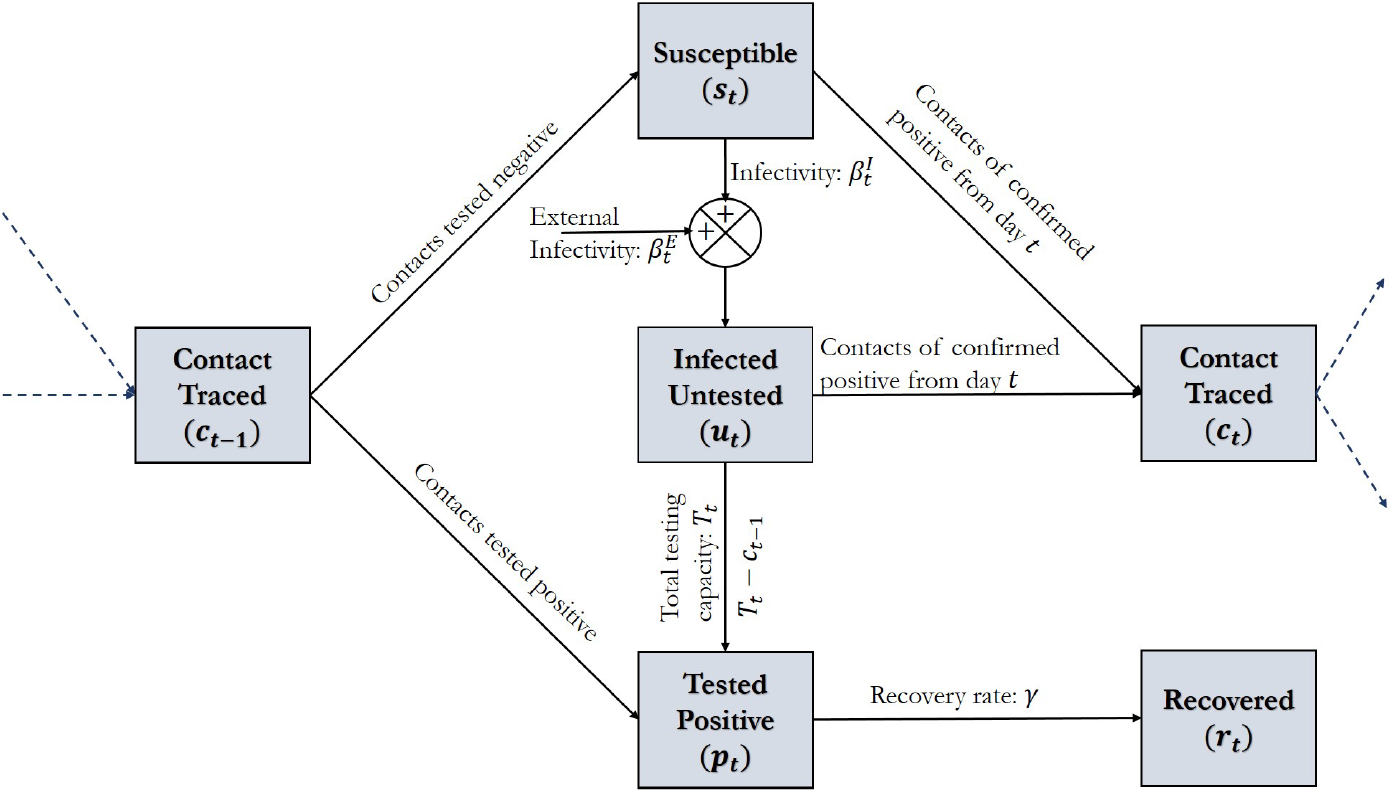
The SUPR Infection Dynamics with Testing and Contact Tracing (where, *s*_*t*_: number of susceptible individuals, *u*_*t*_: number of undetected infected individuals, *p*_*t*_: number of positive infections detected from testing, *r*_*t*_: number of recovered individuals, *c*_*t*_ number of contacts traced on dat *t, T*_*t*_: total number of tests, 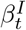: internal infectivity, 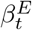: external infectivity, and *γ*: recovery rate.)

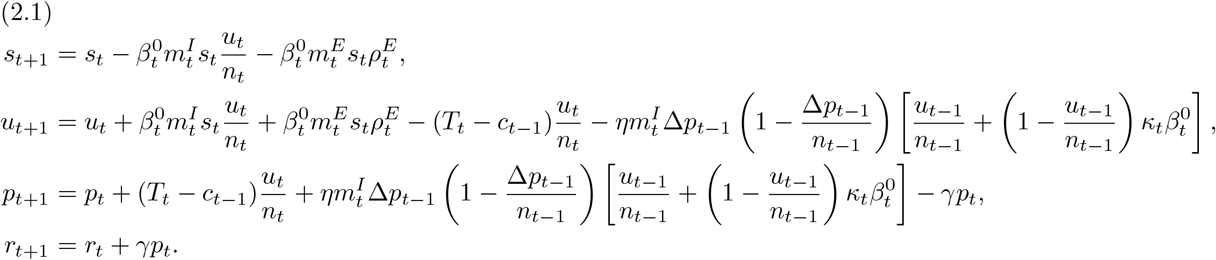

We remark that the above epidemic model lends itself to simplifications when the population is very large. While such simplifications allow for closed form analysis, they are not quite appropriate when the considered population is small as that within an institution. Rather than pursuing such closed-form analysis, here we estimate key parameters of our model from data collected at several universities across the US. These parameter estimates are then plugged in an agent-based simulation that reveals interesting insights into strategies necessary to safely reopen an educational institution.

### 2.2. Parameter Estimation from Data

We estimate a subset of the parameters of our dynamic epidemic model from data. In this section, we describe the data and the parameter estimation techniques used in the empirical and simulation studies.

We utilize the daily number of tests and infections from the SHIELD testing program at the University of Illinois at Urbana-Champaign (UIUC), Champaign County, IL. In addition, we use less granular data of weekly new infections and testing conducted at 85 large universities other than UIUC across 78 counties in the US (see list in Appendix B). While data from more universities are available, we only used universities that conduct some bulk testing for random screening purposes. A non-random testing strategy is unsuitable for providing a credible estimate of institutional infection rates, and hence, do not conform to our modeling. The data on COVID-19 infections at the universities across the US were collected from COVID-19 dashboards maintained by respective universities. For example, the data for UIUC was collected from https://covid19.illinois.edu/on-campus-covid-19-testing-data-dashboard/. The data from tests at these universities is augmented with information about COVID-19 infections in the counties within which the universities are located to estimate parameters pertaining to infections from external contacts. We use the data from the Johns Hopkins Coronavirus Resource Center at https://coronavirus.jhu.edu/. A detailed description of the data collection and data are available in the following website: https://public.tableau.com/profile/anton.ivanov3554, which is managed and maintained by one of the co-authors.

To explain the estimation process, consider the dynamics of the untested infected population *u*_*t*_ in (2.1). Notice that the strength of the untested infected population is not observed, making it difficult to estimate parameters directly from data without making simplifications. In what follows, we justify the simplifications we make using the data from the SHIELD program and then describe our data fitting approach. First, we assume that the segment *p*_*t*_ that has tested positive and is isolated from the rest of the population is relatively small compared to the total population of an institution, i.e., *n*_*t*_ ≈*n* and *s*_*t*_ ≈*n* − *u*_*t*_ − *r*_*t*_. The UIUC data reveals that the typical number of people that test positive tests is <100. If individuals recover roughly in the span of two weeks, we expect the number of quarantined individuals at any particular day to be around 1K-2K, that is much smaller than the total population of 50K in UIUC. We remark that even during the initial surge of infections immediately following the re-opening of UIUC in August 2021, the total isolated individuals on a day was below 5% of the population. Second, we assume that the positivity rate among the daily bulk tests is a good approximation of the infection incidence within the whole population within the institution, i.e., Δ*p*_*t*_*/T*_*t*_ ≈ *u*_*t*_*/n*. This assumption is reasonable within UIUC, given that UIUC is testing almost 20% of its population daily. Third, we simplify contact tracing by using an approximate value for *κ* for parameter estimation. At UIUC, the number of daily tests conducted traced contacts has typically ranged in 50 − 250 that is much smaller than the 10K daily bulk testing. We emphasize that even though we ignore contact tracing in estimating parameters, we do not ignore it in agent-based simulation as we precisely study the role of contact tracing and bulk testing in infection mitigation.

The dynamics of the untested infected population from (2.1) can be translated into the dynamics of the positivity rate *ν*_*t*_ = Δ*p*_*t*_*/n*_*t*_ ≈ *u*_*t*_*/n* based on the aforementioned approximations as

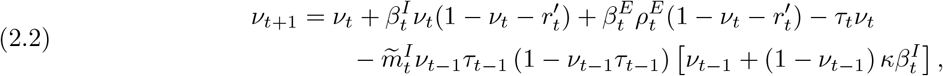

where we have used the notation

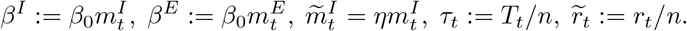

We have the data of total daily tests *T*_*t*_, strength of the recovered group *r*_*t*_, and the daily new detected infections Δ*p*_*t*_ over the course of *D* = 14 weeks from the UIUC SHIELD program and the external infection load 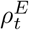 in Champaign county where UIUC is located, over the same period from the Johns Hopkins Coronavirus Resource Center. From the values of *T*_*t*_, *r*_*t*_, Δ*p*_*t*_, we can infer 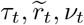 in (2.2). Our goal is to estimate the parameters

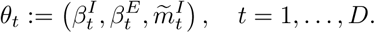

Given *ν*_0_ and the measured values of 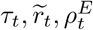 for t = 1, …, *D*, the parameters Θ := (*θ*_*1*_, …, *θ*_*D*_) determine the trajectory of positivity rates *ν*(Θ) over the *D* days, per (2.2). The goal of the estimation process is to find Θ that minimizes an error between the observed trajectory of positivity rates 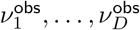 and *ν*_1_(Θ), …, *ν*_*D*_(Θ). To find *θ*_*t*_, define Θ_*t*_ := (*θ*_*t*_, … *θ*_*t*_) that assumes the same value of the parameter for each time over the *D* days and minimizes the weighted squared error

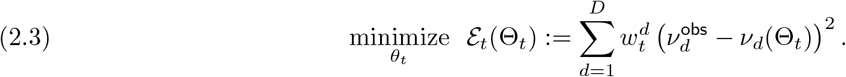

The weights *w*’s are positive and weigh the error between observed positivity rates *ν*^obs^ and the same implied by the parameters *ν*(Θ_*t*_). The weight is more around day *t* and less so as *d* gets further away from *t*. We specifically choose 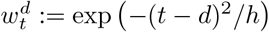, where *h* is a tuning parameter to minimize error in estimation. This estimation process solves *D* nonlinear regression problems to compute the parameters, one for each day. The regression problems are nonlinear as the parameters enter the *ν*-dynamics in (2.2) non-linearly. We remark that such an estimation process can detect variations in the estimated parameters over time and provides us a way of identifying the efficacy of testing and other infection mitigation measures. Finally, from the non-linear regression we obtain the estimates for 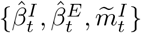, which enables us to estimate rest of the parameters as 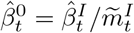 and 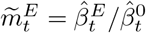.

### 2.3. Agent-Based Simulation

The analytical epidemic model in (2.1) mathematically describes the infection and the testing dynamics. However, this model alone is not sufficient for robust policy evaluations–the goal of our current paper. To appreciate why that is the case, notice that the epidemic model is deterministic and is meant to capture the average infection dynamics through time. The possible deviations from the average will depend on the inherently random interactions among people and disease transmission. The compartmental model cannot adequately capture this randomness. This is the intrinsic downside to using compartmental models that seeks to represent the infection status of *n* individuals (with 4^*n*^ possibilities at any time *t*) through the dynamics of the size of different population segments with respect to the disease. While different policies for testing and other preventative measures can be approximately evaluated based on (2.1), such evaluations are not robust to the randomness of this dynamics. Jaffrey and Treur (2008)^29^ show that for relatively smaller populations, the agent based analysis provides better representation of the infection dynamics than that provided by compartmental models. To address the challenge, we adopt a different strategy in this section. Specifically, we build scenarios of daily interactions among individuals and track the infection status of *n* agents in simulations. Each simulation run of these *n* agents is random and provides one sample path of the infection dynamics through time. Our setup is such that the aggregate dynamics indeed mirrors the mathematical model we presented in (2.1). We also relax several simplifying assumptions made to derive the analytical model in these agent-based simulations. Different testing policies are then evaluated over multiple sample paths with less restrictive assumptions, making the resulting policy evaluations more immune to said assumptions and the randomness of the infection process.

Agent-based simulations have been extensively used in the context of epidemic spreads and transmissions.^29–32^ In Algorithm 1, we outline the steps of our agent-based simulation. In our setup, we create *n* agents who interact with each other and with people outside the institution following simple probabilistic rules. We track the status of *n* individuals. At any given time, their status is one among susceptible (S), undetected infected (U), detected positive (P) or recovered (R). Agents can randomly transition from one state to another based on the random interactions and chances of infections and recovery. The dynamics of Figure 1 guides the transitions for each individual. We maintain the contact list *C* of individuals to simulate the effects of testing via contact tracing.

For the dynamic interactions and disease transmission, we let each agent meet other agents randomly in a way that the average number of contacts within and outside the institution are the average values of *m*^*I*^ and *m*^*E*^, respectively. New infections emerge with a probability *β*^0^. Recall that our analytical model allows for possibly time-varying number of tests *T*_*t*_, mobility parameters 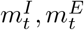 and base infectivity 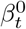. In the agent-based simulations, we choose constant values of these parameters within a simulation run over *D* = 120 days and drop the subscript in their symbols for brevity. These parameters are varied across different simulation runs from among the values obtained from parameter estimation with data over multiple days. With each choice of such parameters, we generate 100 sample paths. We evaluate the outcome of an experiment through the following metric

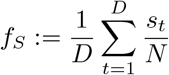

that measures the average number of susceptible individuals as a fraction of the total population. This measure can also be viewed as the area below the trajectory of the number of susceptible people in the institution (area below the susceptibility curve), normalized by the population size. Since *s*_*t*_ ≤ *N*, we have *f*_*S*_∈ [0, 1]. A higher value of *f*_*S*_ indicates a healthier outcome as it implies that less number of people on average are infected. Policy evaluations are made based on both the mean and the range (95% confidence interval) of *f*_*S*_ over *D* = 120 days.

We are able to study the effects of several refinements through agent-based simulations that do not appear in our analytical model. For example, in one of our experiments, we consider two different risk-groups within the population, one with a higher risk of infection transmission due to higher contact rates than another. Other example deviations from the analytical setup includes modeling the effect of delay *δ* between testing and the revelation of test results, imperfect isolation of detected COVID-positive individuals (isolation efficiency *ψ*) and possibilities of false negative tests (test sensitivity *χ*).

## 3. Results and Discussions

First, we report the results from parameter estimation using data from various universities. Then, we present the results from our agent-based simulation with estimated parameters to glean interesting insights into reopening strategies.

### 3.1. Parameter Estimation From Data

We first present the results from the UIUC SHIELD program and then compare these results from those we obtain from other US universities. Figure 2a shows the daily number of COVID-19 tests and the daily new detected positive cases at UIUC. Over the 14 weeks of the Fall 2020 semester, daily tests averaged at 7, 964 with a standard deviation of 3, 525. The large variations in daily tests can be explained by lower testing conducted during the weekends and the adaptive bulk testing policy adopted by UIUC. By adaptive, we mean that the necessary testing frequency has been changed to cope with positivity rates over time. For example, right after the school reopened, the incidence of infection was quite high (between August 1, 2020 and August 25, 2020). Figure 2b captures this increased positivity rate mid-August. Stricter social distancing and mask wearing measures were implemented, which included increased surveillance and enforcement of CDC guidelines for COVID-19. At the same time the frequency of testing was increased from twice a week to thrice a week for students and from once a week to twice a week for faculty and staff.

**Figure 2.**
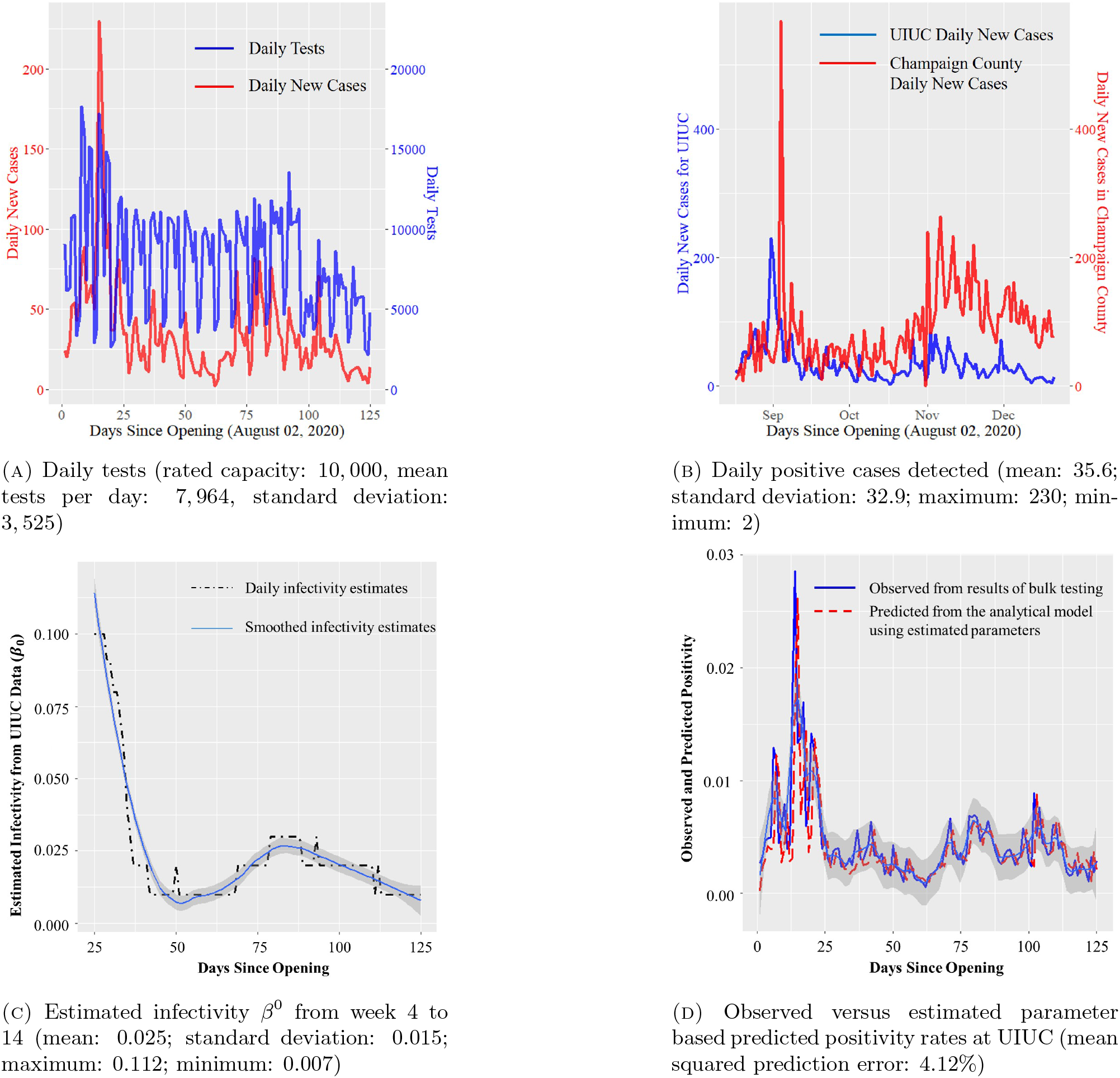
Visualization of the data and the results of parameter estimation from the UIUC SHIELD program.

Table 1 records the results of parameter estimation from the data of the UIUC SHIELD program with 95% confidence intervals. In Figure 2c, we present the weighted least square estimates for base infectivity *β*^0^ starting from the fourth week (day 25) to the end of week 14. Our estimation process yields *β*^0^ for each day, shown as a dashed black line in the figure, with a smoothed variant drawn as the blue line. The average *β*^0^ is 0.025 with a standard deviation of 0.015. The estimates corroborate the higher infection rates around 0.1 after reopening. Several media reports criticized UIUC’s reopening strategy based on this initial high infectivity rate. After the elevated infection rates in the beginning, however, the rates quickly dropped to around 0.01. While the UIUC population took some time to get used to the new reality of operating with

**Table 1.** Parameter estimates from UIUC SHIELD data

| Parameter | Mean | Std. Dev | 95% CI | Range |
| --- | --- | --- | --- | --- |
| $\beta^0$ | 0.025 | 0.015 | 0.01-0.075 | 0.007-0.112 |
| $m^I$ | 5 | 1.35 | 1-6 | 1-11 |
| $m^E$ | 2 | 0.41 | 0-3 | 0-3 |
| $\rho_t$ | 0.043 | 0.022 | 0.012-0.083 | 0.009-0.968 |
| $\tau_t$ | 0.159 | 0.071 | 0.112-0.237 | 0.042-0.351 |

#### Algorithm 1: The Agent-Based Simulation Setup

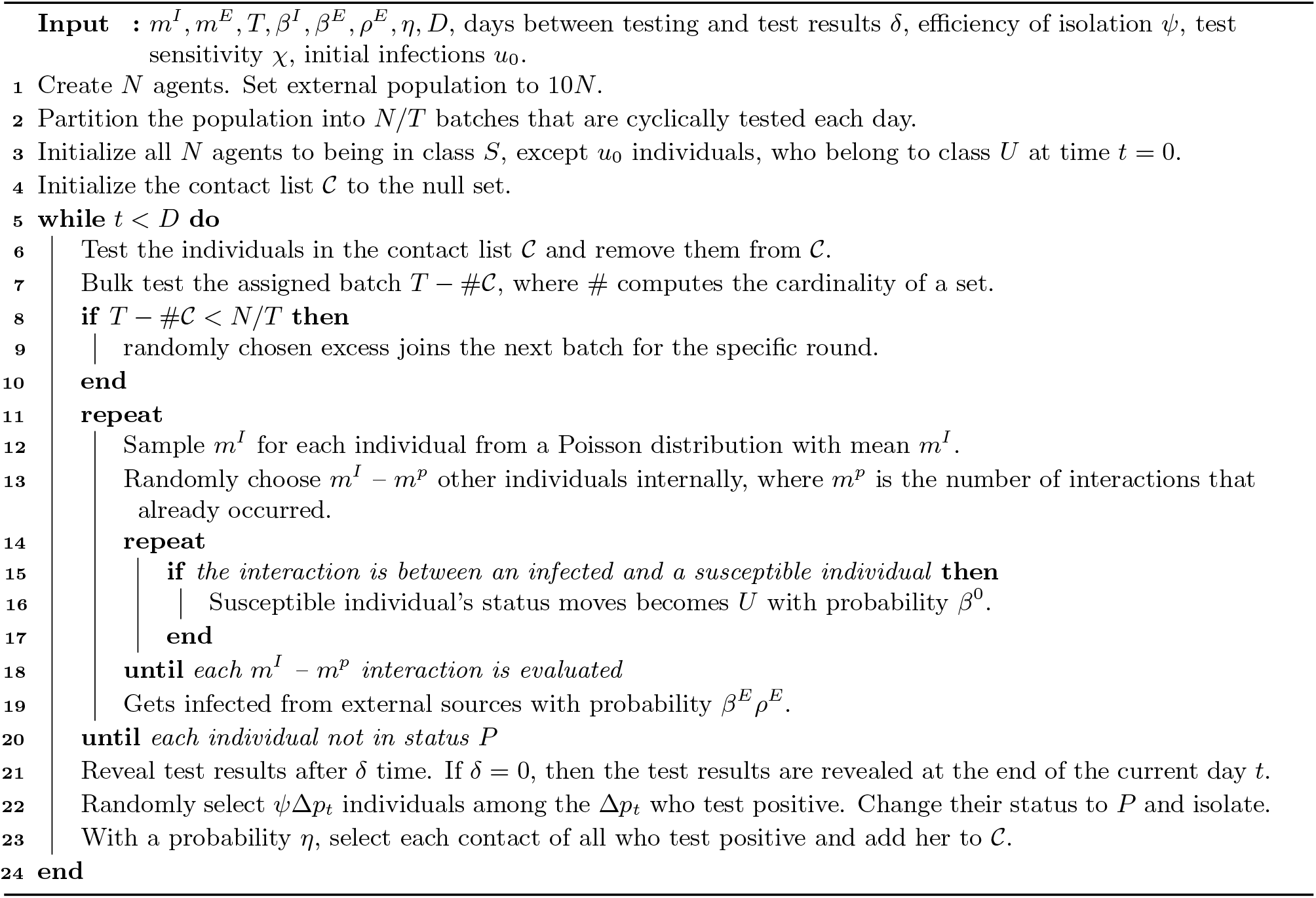

COVID-19, sound policies related to social distancing and mask wearing enforcement rapidly took effect. The climb in infection rates to 0.03 in week 11 (both in Figures 2a and 2b) is explained by the diadic relationship between an institution’s internal infections and the surrounding environment’s infections. In Figure 2b we show the plots of daily new cases for UIUC as well as Champaign county. The figure indicates that the external infections and the internal infections of UIUC are not independent. While the first surge of infections in the initial weeks of reopening is associated with a large inflow of students from outside, the second surge in infections in Champaign county where UIUC is located, and the internal infections from UIUC’s SHIELD program is probably related to the general rise in infections due to several reasons including increase in social contacts due to elections, social unrest and lockdown fatigue in the general population. This second surge in institutional infections is not a characteristics of UIUC alone. Rather, the second rise in infections is observable in all other universities as shown in Figure 3b, which we discuss later.

**Figure 3.**
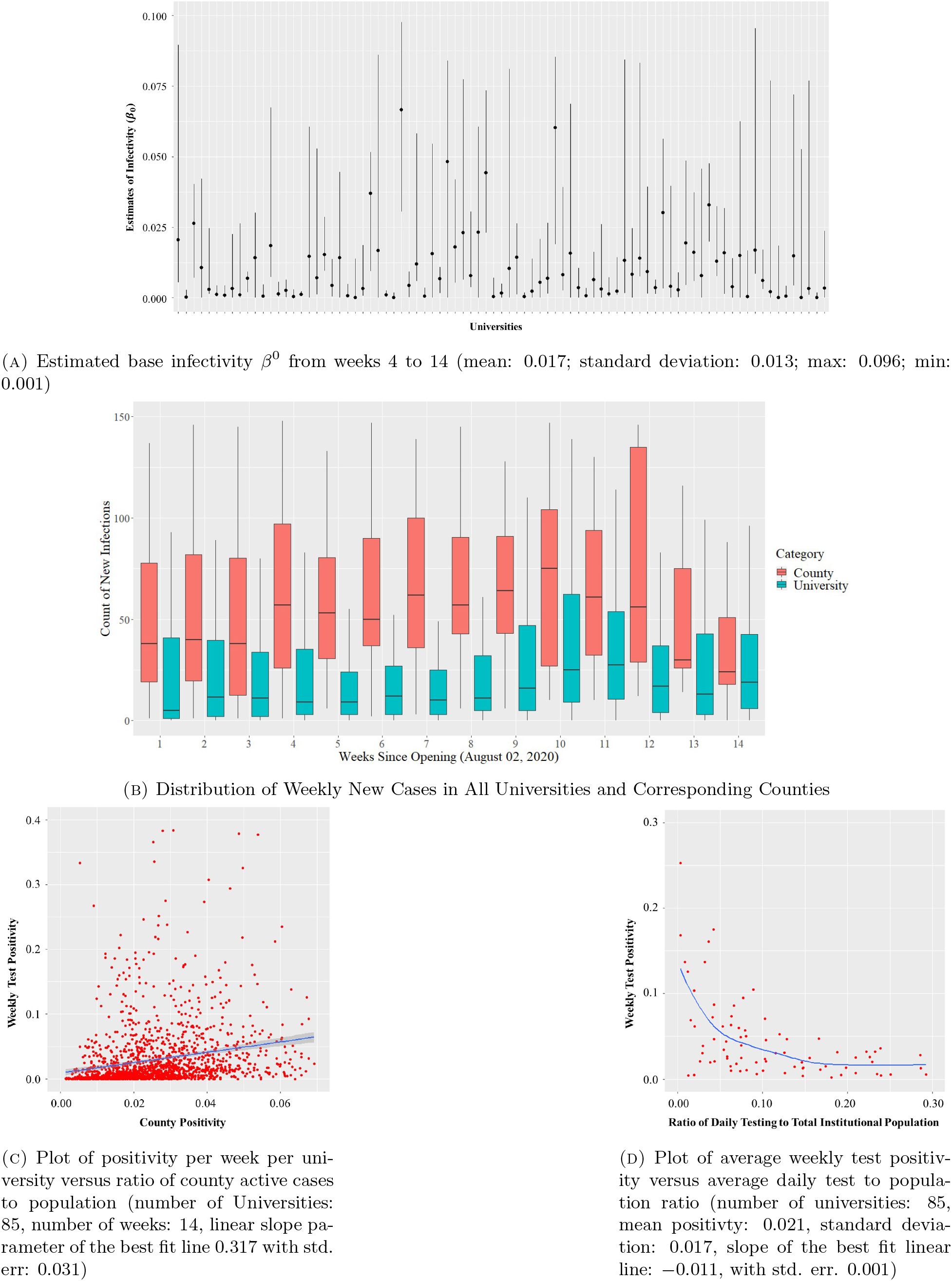
Relevant plots generated using the data from 85 universities in the US (see Appendix B for the list).

The estimated contact rate *m*^*I*^ within UIUC is 4.85≈ 5 with a range of [1, 11] and standard deviation of 1.35. The median contact rate from the estimates is 3. Similarly, the mean and median external contact rate *m*^*E*^ is 2. While *m*^*I*^ > *m*^*E*^, the proximity among these estimates indicate that the prevalence of infection in the county around the institution contributes heavily towards the infections within the institution. Recall that *β*^*I*^ = *β*^0^*m*^*I*^ denotes the internal infectivity rate. This rate is estimated at 0.12 with a standard deviation of 0.02. One often uses the basic reproduction number of an epidemic as a metric of how fast an epidemic is growing. This number is given by *R*^0^ = *β*^*I*^*/γ*, the ratio of the infection rate within the institution and the recovery rate. For UIUC, *R*^0^ is estimated to be 1.82 with a 95% confidence interval of 0.75 − 3.05 for an average recovery period of 15 days. Early published estimates put *R*^0^ for COVID-19 in the range 3.40 −3.67.^16, 33^ Thus, our estimates for UIUC are significantly lower than published estimates. We suspect that the difference between these estimates emanates from the institutional setup that is significantly different than that of general social life. The differences arise in terms of the population sizes, the ability of institutions to enforce preventative measures such as social distancing, mask wearing and extensive facility sanitization.

To validate our estimation, we computed one day look-ahead prediction of the positivity rates *ν*_*t*_ using the estimated parameters and compared these predictions to observed positivity rates. Figure 2d shows that the day-ahead estimates indeed match well with observed data (with a mean prediction error of 4.12%).

Table 2 shows the results of parameter estimation using weekly testing and infection data from 85 universities across the US other than UIUC; the list is included in Appendix B. Figure 3a shows the estimates of *β*^0^ with an average of 0.017 across all 85 universities. The internal and external mobility estimates *m*^*I*^ and *m*^*E*^ are 3 and 1, respectively. While these numbers vary across universities and vary over time within universities, they are quite similar to those obtained for UIUC. The parameter estimates from the weekly data from these universities together with those from the UIUC SHIELD program provide us with a range of parameters for the agent-based simulation later in this section.

**Table 2.** Parameter estimates using data from 85 US universities other than UIUC.

| Parameter | Mean | Std. Dev | 95% CI | Range |
| --- | --- | --- | --- | --- |
| $\beta^0$ | 0.017 | 0.013 | 0.005-0.067 | 0.001-0.096 |
| $m^I$ | 3 | 1.02 | 1-5 | 1-5 |
| $m^E$ | 1 | 0.28 | 0-2 | 0-3 |
| $\rho_t$ | 0.025 | 0.016 | 0.002-0.057 | 0.001-0.127 |
| $\tau_t$ | 0.083 | 0.104 | 0.006-0.750 | 0.004-0.300 |

In Figure 3b, we include a box-plot of the total number of infections in the universities and in the counties where the universities are located. Notice that the infection count within the universities and the counties both show a surge in the month of November. The UIUC data shows a similar surge in Figure 2b. This elevated infection count is possibly a result of the confluence of multiple factors that include US elections, the rise in socio-political uprisings and COVID-19 lock-down fatigue among the general populace. Another important factor that may have resulted in the overall increase in infections in the environment is dropping ambient temperature due to the approaching winter season.^34, 35^ In our analysis, we do not include the effect of climate due to lack of data and due to the fact that all universities that we analyze are located in the United States, and therefore, the climate variation is not too high. This is a simplifying assumption that we have used for our analysis, and therefore, is a potential limitation. Motivated by the similarity in the variation of the infection counts within and outside the university, we plot the weekly test positivity for all 85 universities against the external positivity of the surrounding environment. The external COVID-positivity is measured as the ratio of the total number of active cases in the neighboring county and the total population of said county. The plot demonstrates a clear positive correlation–a linear fit yields a slope of +0.317 with a standard error of 0.031 (p-value: < 10^−16^). This plot illustrates that incidence of COVID-19 infections within an institution affects and is affected by the infections in the neighboring counties. This data analysis validates our modeling choice to include external infection load *ρ*_*t*_ and external contact rate *m*^*E*^ in the dynamical system model for the epidemic in (2.1).

Through the agent-based simulation later in the section, we argue that rapid bulk testing is key to safely reopening educational institutions. Before we present the results from our simulations, we remark that weekly COVID-19 positivity rates from these 85 US universities indeed exhibit negative correlation with the extent of testing conducted at the universities. See Figure 3d for the plot of the positivity rates against the ratio of the daily tests conducted at the universities to the institutional population.

### 3.2. Agent-Based Simulation To Evaluate Reopening Strategies

We now report the results from agent-based simulations to understand the effects of various parameters such as the extent of bulk testing, efficiency of contact tracing, preventative measures that reduce base infectivity, etc. on the dynamics of the infection process. Majority of the results utilize parameter estimates from the UIUC SHIELD program with *n* = 50K. The parameters are chosen from the range obtained from the estimation step for each of the 100 runs. These parameters, however, are held constant through *t* = 1, …, *D* in each run, unless otherwise specified.

We remark that we have conducted upwards of a million simulations with different combinations of parameters, over and beyond what we report here. As a result, we believe our policy evaluations to be robust and useful for practical policy guidelines. However, we do not claim optimality of our guidelines in a statistical sense and leave such a quest to a future endeavor.

#### 3.2.1. Bulk testing capacity

With the parameters *β*^0^ = 0.025, *m*^*I*^ = 5, *m*^*E*^ = 2, and *ρ*^*E*^ = 0.043 estimated from the UIUC data, we simulated four different scenarios of bulk testing with constant daily tests of ∈ *T* {1K, 5K, 10K, 15K} over a period of *D* = 120 days roughly spanning a semester. In our simulations, we assumed the efficiency of contact testing to be *η* = 0.9 and the efficiency of isolation to be *ψ* = 0.95. To make our simulations more realistic we assumed that the tests have a sensitivity of 0.92. See Wyllie at al. (2020)^22, 36^ that reports the sensitivity of saliva-based LAMP tests to be between 0.90 and 0.95. Given that the average delay between conducting a test and revealing the test result at UIUC is generally below 12 hours, we assume that test results are available immediately in our simulations. In Figure 4a, we plot the size of the susceptible population across time in our simulations. For all experiments, we set the initial number of infection *u*_0_ = 5. One of our test runs with 10K daily tests lead to a total of 11, 041 infections in a span of 4 months. This number is close to 10, 890 infections that we obtain from simulating the analytical model in (2.1) with 10K daily tests–a step that verifies that the analytical model and the agent-based simulations are consistent. The agent-based simulation, however, is much more powerful for policy design as it captures the stochastic nature of the infection dynamics that permits robust policy evaluation.

**Figure 4.**
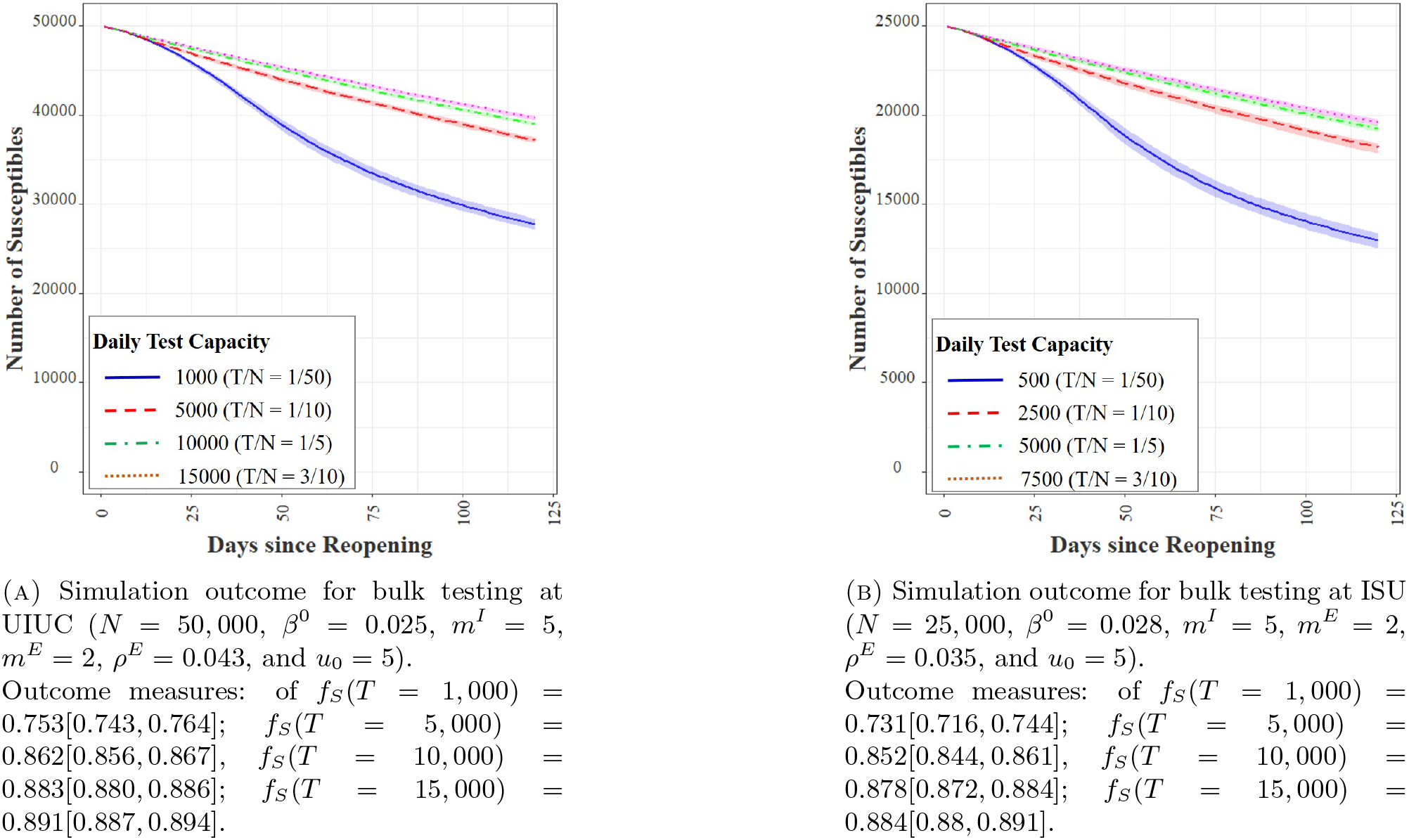
Comparison of the effect of bulk testing at UIUC and ISU.

The simulations reveal that the marginal benefits of testing capacity is high at lower testing capacities. For example, moving daily tests from 1K to 5K reduces the average fraction of total infected from 0.247 to 0.138, a reduction of 44%. This translates to a total of 5, 450 less infections over a span of 120 days. However, increasing the capacity from 5K to 10K daily tests reduces the same fraction to 0.117, a reduction of only 15%. Increasing daily testing to 15K decreases the same to 0.109, a reduction of 7% from 10K tests per day. Without bulk testing, we obtain *f*_*S*_ = 0.710, which indicates that the total infection is 2.48 times higher than that obtained with 10K daily tests. This translates to a total of 8, 650 less infections over 120 days that result from bulk testing at the rate of 10, 000 daily tests on an average. In other words, bulk testing can dramatically reduce the number of infections and should ideally form a central component of reopening strategies for educations institutions.

We perform a similar analysis for Illinois State University (ISU) for which the parameters of infectivity and contact rates are similar to that of UIUC, but its institutional population is around half of that of UIUC.

The similarity between the outcomes in Figure 4a for UIUC and in Figure 4b for ISU demonstrates that *τ* = *T/n*–the ratio of daily tests to the population size–plays a determining role on the infection dynamics. In the simulations described above, we have fixed the daily number of tests throughout the *D* days. In practice, operating with a fixed capacity is typically inefficient and may lead to higher costs compared to adaptive testing capacities. UIUC has undertaken an adaptive approach to testing. For example, upon opening in the beginning of August 2020, UIUC required all students to get tested once a week. From August 16, the university mandated all students and faculty/staff to test twice a week due to increased positivity within the university. On September 9, the requirement for faculty and staff was dropped to once per week, following the dampened rate of infections within the university. On November 2, the requirement was ramped up to thrice a week for students and twice a week for faculty and staff as positivity rates increased both within and outside UIUC. In view of the above, we seek to understand the effect of adaptive testing policy. Therefore, we perform an agent-based simulation where on each day *t*, the testing capacity *T*_*t*+1_ for the next day grows with the ratio of the positivity rates on day *t* and *t* − 1. By positivity rate on a day, we mean the ratio of number of positive infections detected to the number of tests conducted on that day. Figure 5 illustrates the result of this experiment. Notice that the average number of daily tests in the adaptive approach dropped to 9,688, that is lower than 10K by 312 tests every day. Yet, average *f*_*S*_ with adaptive testing capacity is 0.887, which is higher than the area under the susceptible curve (0.883) obtained with 10K daily tests. Recall that rapid saliva testing costs $20 - $30 per test. Thus, adaptive testing leads to an estimated cost saving of $7.5 - $1.1 million over *D* =120 days, while performing better on the disease mitigation front than fixed testing capacity. In this simulation, the daily tests fluctuate significantly during the initial periods, which dampen after the infection load stabilizes. In practice, however, such daily fluctuations may be difficult to administer, requiring a smoother variation in testing policy similar to that adopted at UIUC.

**Figure 5.**
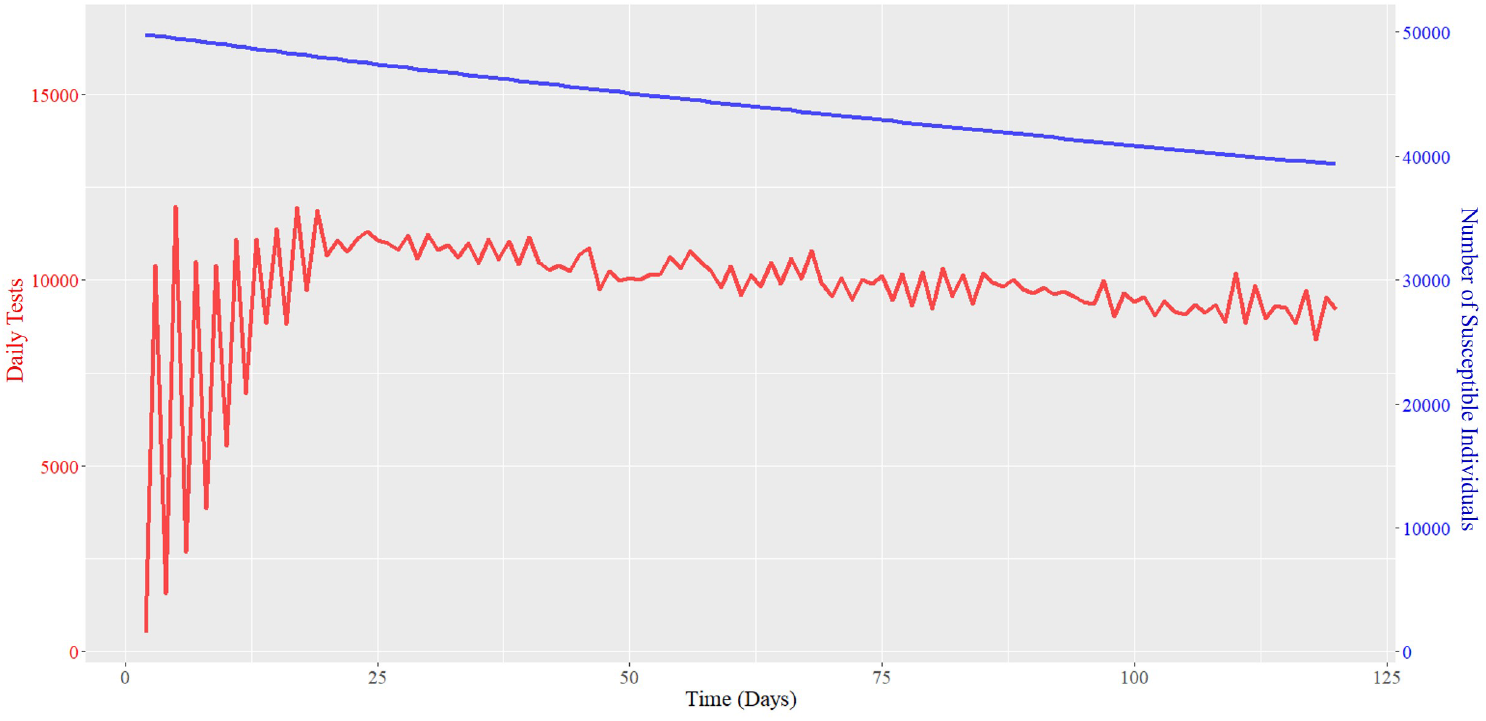
Adaptive testing outcome with maximum test capacity of 15, 000 (average daily tests: 9,688, maximum daily tests: 11,998, *f*_*S*_ : 0.887

#### 3.2.2. Efficiency of contact tracing

Efficiency of contact tracing is understood as the probability with which a contact of an infected positive individual is identified and tested. We report our empirical findings for contact tracing efficiencies of 90% and 80% in Table 3. The results indicate that contact tracing efficiency has much more impact on the epidemic dynamics when bulk testing capabilities are small. This impact almost disappears when bulk testing capabilities increase. For example, with bulk testing 1K individuals daily, contact tracing efficiency drop from 90% to 80% leads to a drop of mean *f*_*S*_ from 0.753 to 0.712 (5.4% reduction). The same numbers with 15K daily tests are 0.891 and 0.890, respectively. While contact tracing helps, our results yield that bulk testing has a much larger impact. With around 10K daily tests with parameters for UIUC, we typically found the number of contacts of positive individuals *c*_*t*_≈ 650 on an average, and with a probability of infection slightly higher (factor of *κ*) than that of random selection approximately 20 positive cases are detected. As a result, the total number of infections detected via contact tracing is much smaller as compared to about 200+ COVID-positive individuals detected via bulk testing.

**Table 3.** Impact of contact tracing efficiency on mean and range of *f*_*S*_.

| $T$ | Contact tracing efficiency $\eta$ | |
| --- | --- | --- |
|  | 90% | 80% |
| 1K | 0.753 [0.743, 0.764] | 0.712 [0.697, 0.724] |
| 5K | 0.862 [0.856, 0.867] | 0.859 [0.854, 0.867] |
| 10K | 0.883 [0.880, 0.886] | 0.880 [0.878, 0.883] |
| 15K | 0.891 [0.887, 0.894] | 0.890 [0.887, 0.893] |

Judging based on our experiments, we find it unlikely for contact tracing alone to define a viable infection containment strategy, given the large proportion of asymptomatic carriers of COVID-19.

#### 3.2.3. Base infectivity and preventative measures

Universities have adopted several measures that directly impact the base infectivity levels, such as mask wearing and frequent sanitization of its premises. Some institutions have even pursued punitive measures for violation of mask wearing measures such as financial penalty, sanctions, and restrictions on accessing institution facilities. For example, at UIUC, several students were placed under probation for violation of regulations related to COVID-19 measures after the initial surge of infections immediately following reopening in August. At UIUC, our estimation puts *β*^0^ in the range 0.01 − 0.11, with a mean of 0.025. We simulate the effect of adopting less stringent preventative measures and report the results of agent-based simulations with *β*^0^ ∈ {0.025, 0.040, 0.055, 0.070} for multiple levels of testing *T*. We plot the outcomes in Figure 6. Interestingly, Figure 6a reveals that with 1K daily tests, the entire population will get infected within 50 days for *β*^0^ ≥ 0.04. Similar catastrophic results ensue even with higher testing capacities (see Figures 6b - 6d) at high values of *β*^0^’s. The impact of *β*^0^ on the infection dynamics is rather pronounced, underscoring the importance of preventative measures. This sensitivity to *β*^0^ is not surprising, given that *β*^0^ directly changes the potency of each meeting between a susceptible and an infected individual. The consequence of each new infection then accumulates fast, given the nature of the epidemic dynamics. Besides bulk testing, it is thus imperative for institutions to enforce mask wearing, place hand sanitizers at various locations, periodically clean classrooms and laboratories, etc. This same sentiment is resonated in existing literature.^37^

**Figure 6.**
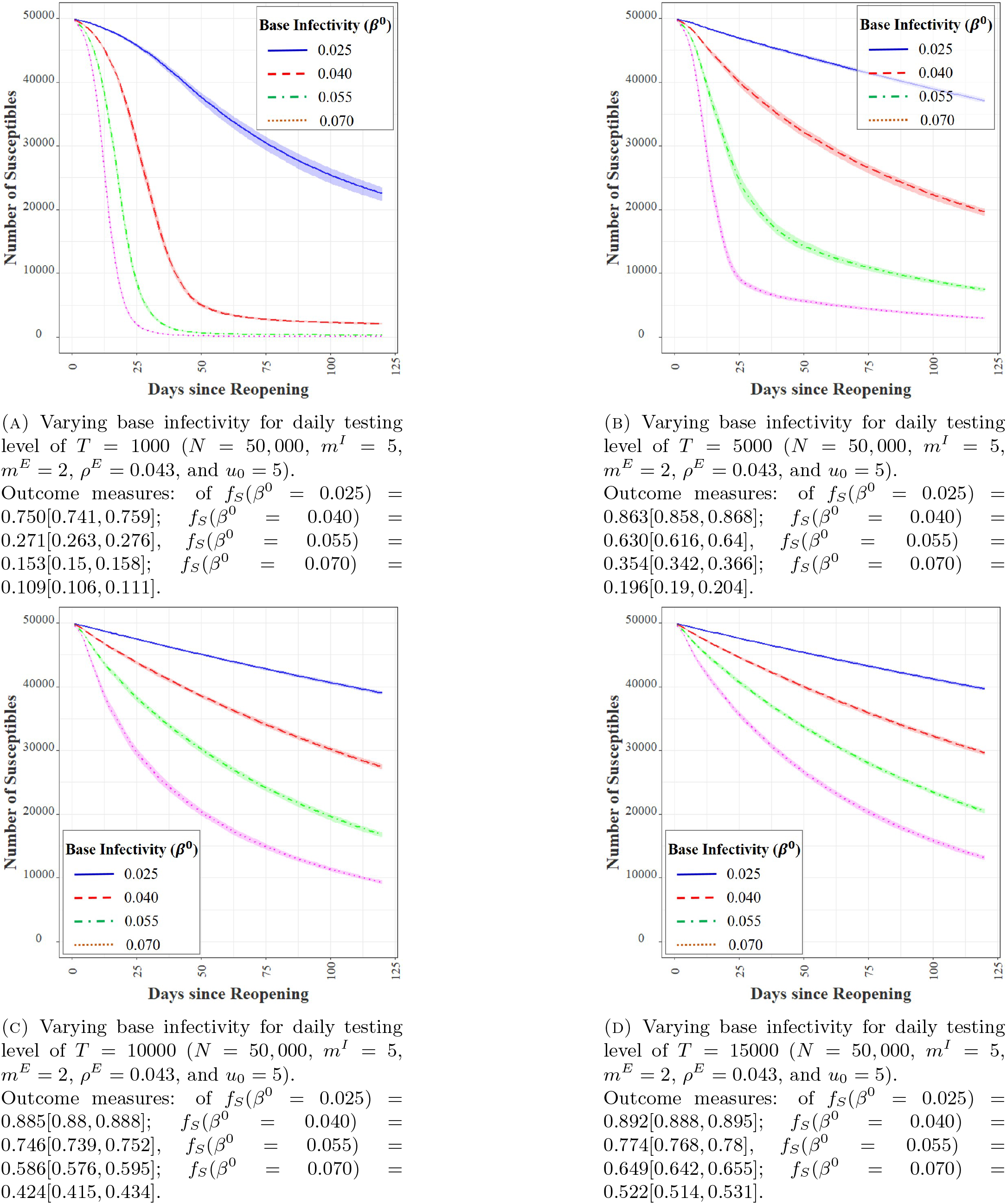
Effect of base infectivity rates on agent-based simulations.

#### 3.2.4. Contact rates

Contacts create opportunities for infection transmission. With the parameters for UIUC (where average *m*^*I*^ is 5 with a range 1 −15), we evaluate the effect of varying *m*^*I*^ from 2 to 11 in steps of 3 in Figure 7. Increasing internal contact rate severely impacts the transmission of infection with testing capacities of 1K and 5K per day. The impact, however, becomes minimal with higher daily testing capacities of 10K and 15K. Strategies to reduce internal contacts include spacing out classroom sitting arrangements, staggering class and meeting times, using larger capacity rooms for classes and meetings, and adopting a hybrid of online and in-person operations as feasible. Our experiments demonstrate that increased bulk testing decreases the need for severely restricting internal contacts, revealing that contact restrictions and testing play a complimentary role in infection mitigation.

**Figure 7.**
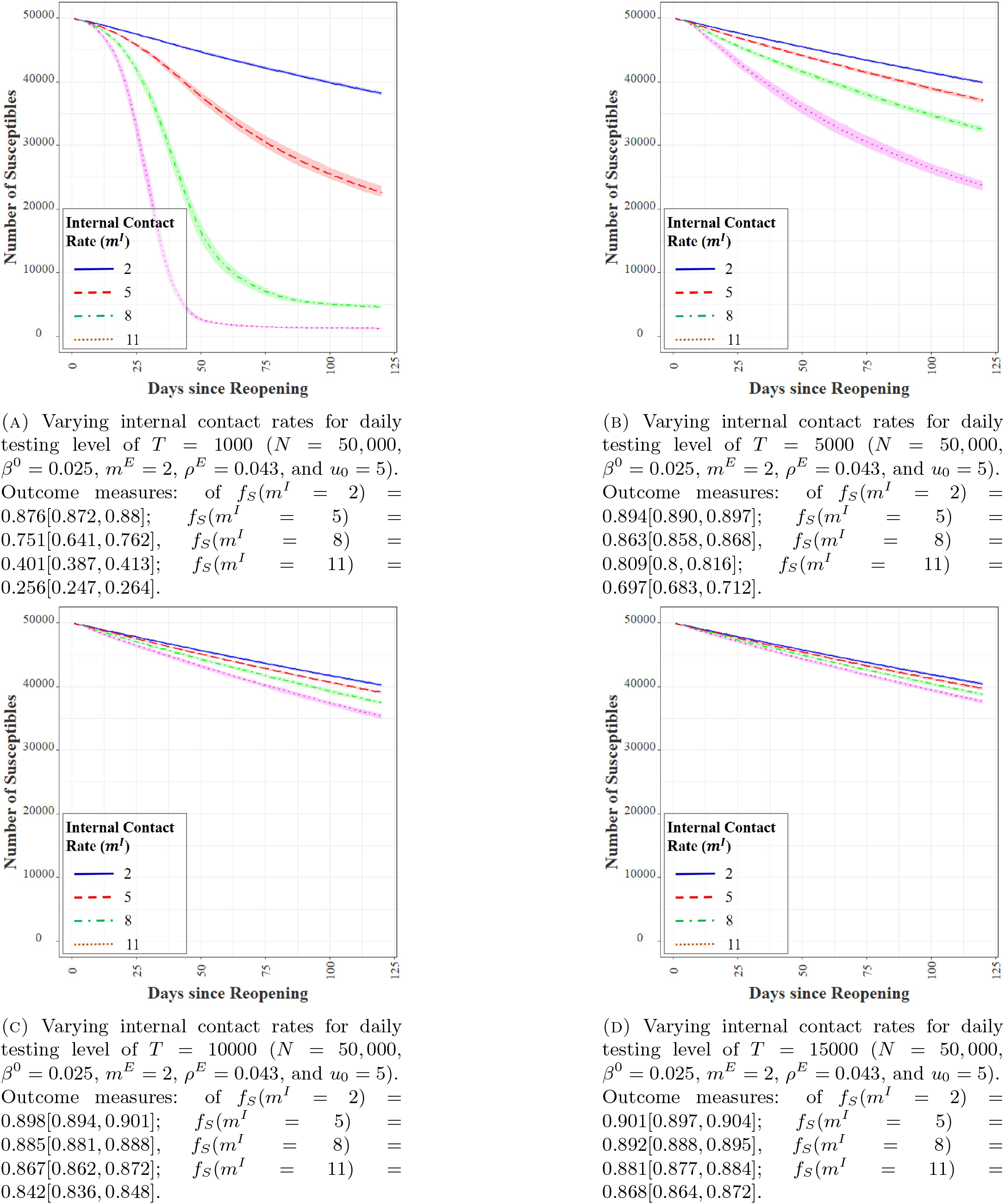
Effect of internal contact rates on agent-based simulations.

The effect of the number of external contacts *m*^*E*^ is similar and the results are omitted for brevity. While an institution may not possess the means to directly control *m*^*E*^, targeted information and awareness campaigns can indirectly reduce *m*^*E*^ by educating the members of the consequences of infection transmission.

#### 3.2.5. Varying testing frequencies among sub-populations

The agent-based simulation results presented so far assume that the institution has a population with homogenous mobilities that we estimate from data. In practice, student groups and faculty/staff typically have different mobilities and hence, belong to different risk categories in terms of their potencies to transmit the disease. Personal communication with the UIUC SHIELD program indicates that they expect the contact rates among the student population to be at least double that of faculty and staff. Based on these expectations, the program has delineated different guidelines for these population groups. Specifically, students were asked to test at least twice a week and the faculty and staff to test once a week over initially, which moved to thrice a week testing for students and twice a week testing for staff and faculty on November 2, 2020 due to increased positivity. Here, we study the impact of risk-based modulation of bulk-testing frequencies through agent-based simulations. To that end, we divide the population of 50K agents in the simulation into two groups–40K students and 10K faculty/staff. We assume that students have an internal contact rate of *m*^*I*^ = 5.5, compared to that of *m*^*I*^ = 3 for faculty/staff.

The numbers are chosen such that the average *m*^*I*^ becomes 5, that approximately equals the rate we estimated from data. Students are then tested at double the rate compared to the faculty/staff. Table 4 presents the simulation outcomes.

**Table 4.** Effect of uniform and risk-based testing frequency on mean and range of *f*_*S*_.

| $T$ | Uniform Frequency | Risk-Based Frequency |
| --- | --- | --- |
| 1K | 0.753 [0.743, 0.764] | 0.784 [0.767, 0.791] |
| 5K | 0.862 [0.856, 0.867] | 0.877 [0.871, 0.884] |
| 10K | 0.883 [0.880, 0.886] | 0.890 [0.887, 0.893] |
| 15K | 0.891 [0.887, 0.894] | 0.892 [0.889, 0.894] |

Compared to the uniform testing frequency, the targeted risk-based testing indeed reduces the overall infection load. The gain from modulation of the testing frequency among the population is higher when the testing capacity is especially limited. For example, the increase in the mean value of *f*_*S*_ is 4.24% (from 0.753 for uniform testing to 0.784 for risk-based testing) with a daily testing level of 1K. The corresponding increase with 10K daily tests reduces to 0.79% (from 0.883 for uniform testing to 0.890 for risk-based testing). Our experiments affirm that targeted testing among the group with a higher mobility (and hence, higher chances of infection) will lead to faster identification and isolation of more COVID-positive individuals, leading to higher values of *f*_*S*_. Such a strategy is especially useful during the initial stages of the infection when testing infrastructure is likely to be limited. While we have only studied two risk classes, a more nuanced risk-stratification of the population can lead to further reductions in infection loads.

#### 3.2.6. Efficiency in isolating COVID-positive patients

While we have so far assumed that isolation is 100%, in reality isolation efficiency tends to vary significantly. For example in China, it was found that 75%-80% of all clustered infections occurred within family. Therefore, in many countries such as in China, South Korea and Singapore COVID-19 patients were isolated in separate facilities rather than at home.^38–40^ In the context of an institution such as UIUC, creation of separate isolation facilities provides high isolation efficiency,^41^ however, isolation efficiency may vary depending on adherence behavior of infected and non-infected individuals. Also, testing is an effective strategy to mitigate infection transmission only if positive detection is followed by proper isolation measures. Here, we study the impact of varying degrees of isolation efficiency *ψ* through our agent-based simulations. This efficiency captures the probability that an individual who tests positive in fact isolates. Table 5 shows the average daily fraction of the susceptible population over 120 days for *ψ* = 100%, 90%, 70% and 50%. The efficacy of testing drops sharply with isolation efficiency and the impact is more pronounced when the number of daily tests is low (see the case with *T* = 1K). Increased volumes of bulk testing can offset the inefficiencies of isolation in part, but that comes at higher costs of building the testing infrastructure.

**Table 5.**
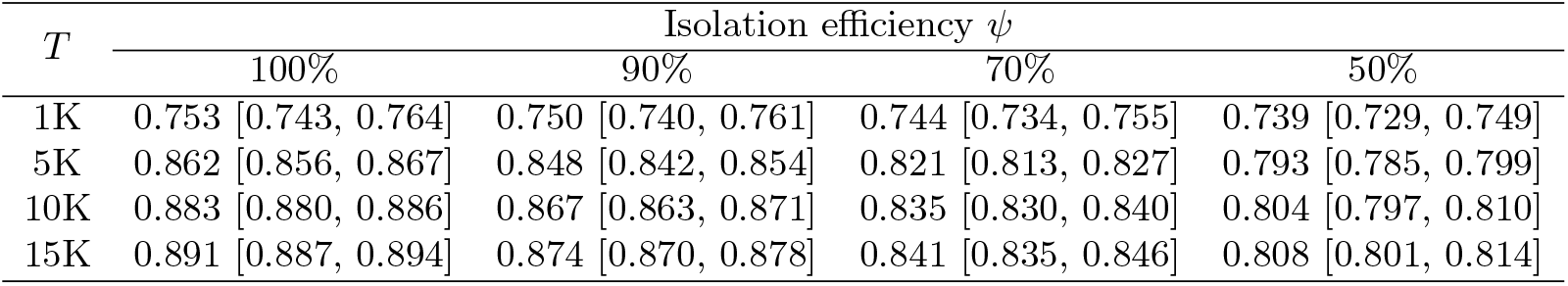
Effect of isolation efficiency on the mean and range of *f*_*S*_.

#### 3.2.7. Delay in obtaining test results

Delay in receiving test results, either due to the nature of testing or due to limited testing capacity as compared to the demand for testing, can have adverse effect on the infections within an institution. In Table 6, we record *f*_*S*_ from our experiments with delays *δ* varied from zero to 4 days in steps of 2 days. The case with *δ* = 0 days corresponds to the setting we considered so far, which is in line with rapid saliva testing at UIUC, where the test results are often made available within 12 hours of testing. As our experiments demonstrate, delay in revelation of test results has a significant impact on the efficacy of testing, even when number of daily tests are high. This is not surprising, given that delay in isolation of infected individuals renders the test somewhat ineffective if these individuals continue to interact with people, awaiting test results.

**Table 6.** Impact of gap *δ* between testing and availability of test results on the mean and range of *f*_*S*_.

| $T$ | Gap between testing and results $\delta$ | | | | | |
| --- | --- | --- | --- | --- | --- | --- |
|  | 0 days |  | 2 days |  | 4 days |  |
| 1K | 0.753 | [0.743, 0.764] | 0.714 | [0.704, 0.724] | 0.698 | [0.685, 0.713] |
| 5K | 0.862 | [0.856, 0.867] | 0.844 | [0.838, 0.850] | 0.823 | [0.817, 0.830] |
| 10K | 0.883 | [0.880, 0.886] | 0.867 | [0.863, 0.872] | 0.849 | [0.843, 0.854] |
| 15K | 0.891 | [0.887, 0.894] | 0.875 | [0.870, 0.880] | 0.857 | [0.851, 0.862] |

#### 3.2.6. Test sensitivities

Our final study seeks to understand the impact of the sensitivity of tests on the infection mitigation strategy. Early reports^22, 36^ claim saliva-based LAMP tests to have an average sensitivity of 92%, i.e., they are able to correctly detect 92% of the cases that are COVID-positive. In contrast, some other reports^19, 20^ show that under certain conditions, particularly with different duration of infections, the test sensitivity can vary widely, and nasal swab based RT-qPCR tests tend to demonstrate much superiod accuracy than the saliva based tests. While we consider bulk testing within institutions, where each individual gets tested relatively frequently (once to twice per week), and the duration of infections may not have a as high a variation as in the case of the general population, yet, we check for sensitivity of bulk testing and isolation policies to varying test sensitivities. In Table 7, we present the outcomes of agent-based simulations with test sensitivities in {90%, 80%, 70%, 60%} with varying degrees of time delays between testing and reporting of test results. All experiments for this study utilized *T* = 10K daily tests. While both the rate of false negatives of the tests and said time delay have adverse effects, the latter appears to be the dominant factor. Higher sensitivity of tests is desirable, no doubt. Even if that efficiency drops, *rapid* bulk testing appears crucial to effectively control the infection growth within the institution.

**Table 7.** Impact of the rate of false negatives together with the the gap between testing and availability of test results on the mean and range of *f*_*S*_ with *T* = 10K daily tests.

| Test Sensitivity | Gap between testing and results $\delta$ | | | | | |
| --- | --- | --- | --- | --- | --- | --- |
|  | 0 days |  | 2 days |  | 4 days |  |
| 90% | 0.881 | [0.875, 0.886] | 0.866 | [0.861, 0.871] | 0.848 | [0.843, 0.852] |
| 80% | 0.877 | [0.873, 0.882] | 0.862 | [0.857, 0.866] | 0.844 | [0.838, 0.850] |
| 70% | 0.872 | [0.867, 0.877] | 0.857 | [0.851, 0.862] | 0.839 | [0.834, 0.844] |
| 60% | 0.865 | [0.859, 0.871] | 0.852 | [0.846, 0.857] | 0.834 | [0.828, 0.839] |

## 4. Conclusions

The reopening of institutions during the COVID-19 pandemic is challenging. The initial experience of reopening in August and September 2020 demonstrate that reopening requires careful planning and measures to mitigate rapid infection spread within an institution. Per a recent media report, several universities have clocked more than 500 cases, such as the University of Alabama at Birmingham (972 cases), the University of North Carolina at Chapel Hill (835 cases), University of Central Florida (727 cases), Auburn University in Alabama (557 cases), Texas A&M University (500 cases), University of Notre Dame (473 cases), and the University of Illinois at Urbana-Champaign (448 cases) within days or weeks of reopening. Our work is motivated to answer if there is any possible policy path that allows institutions to manage the disease, if not fully stop it.

To study epidemic mitigation strategies, we first formulated a dynamical system model to describe the spread of COVID-19 within an institution. The key features of this model include the asymptomatic transmission of the disease, the effect of two channels of testing (contact tracing and bulk testing) and subsequent isolation of those who test positive. The analytical model is parameterized. We used COVID-19 data from 86 universities in the US (including that from the UIUC SHIELD program) to estimate some of these parameters via non-linear regression. The range of parameters were utilized as inputs to an agent-based simulations setup. The outcomes of this simulation are sample paths of the epidemic within the institution. The mean and the range of the outcomes helped us to derive important insights into the efficacy of various parameters and reopening strategies. Having grounded our study to the context of the UIUC SHIELD program data and cross-validated with data from 85 other universities, we believe that our observations are fairly robust and suitable to guide policies at educational institutions.

Our study yields three key observations. First, preventative measures such as mask wearing, social distancing and reduction of contact rates among individuals are indispensable to even consider reopening. Such measures are vital to reduce the potency of asymptomatic transmission. Second, contact tracing is not enough to contain the infection spread. Even though testing infrastructure is expensive, bulk testing capabilities are crucial to contain the disease. The key design parameter is the ratio of the total number of daily tests to the institution population. Additional measures can help combat the disease propagation such as increasing testing frequencies for subgroups with higher mobilities and increasing the efficiency of isolation of patients who test positive. Third, the testing technology should be able to provide test results quickly. The rapidity of the testing cycle appears even more important than test sensitivity (within reasonable limits). Therefore, institutions considering reopening must invest in COVID-testing for its members that is cost-effective, easy to administer in high volumes, and has a quick turnaround time to results.

## Data Availability

The simulation codes will be made available on request.

## Appendix A. Steps In The Derivation of The Analytical Epidemic Model

### A.1. Additional notations used for the derivation

Define the following:

- 𝒮_*t*_ as the set of individuals who are susceptible;
- 𝒰_*t*_ as the set of individuals who are infected but undetected;
- 𝒫_*t*_ as the set of individuals who are infected and detected;
- ℛ_*t*_ as the set of individuals who are recovered;
- 𝒩_*t*_ is the set of mobile individuals who are not isolated, i.e., 𝒩_*t*_ = 𝒮_*t*_ ∪ 𝒰_*t*_ ∪ ℛ_*t*_;
- 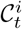is the set of contacts of individual *i* at time *t*;
- 𝒞_*t*_ is the set of all contacts in the contact list for day *t* to be tested in day *t* + 1;
- Δ𝒫_*t*_ is the set of new detected positive individuals on day *t*;
- *x* → *y* as *x* meets (comes in contact with) *y*;
- # as the cardinality of a set such that #𝒰_*t*_ = *u*_*t*_;
- 𝒜 = {*x* : *x* ∈ 𝒳}, an arbitrary a set 𝒜 of elements of type *x* belonging to a super-set 𝒳;
- 𝒳 ∨ 𝒜; 𝒴, is the union of set 𝒳 and set 𝒴;
- 𝒳 ∩ 𝒴, is the intersection of set 𝒳 and set 𝒴;
- *i, j, k* are individual members of an institution.

### A.2 Derivation of the number of new infections

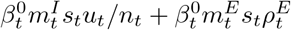. At time *t*, the number of infected undetected individuals are *u*_*t*_, the number of susceptible individuals are *s*_*t*_, and the number of recovered individuals are *r*_*t*_. The infected and detected individuals, *p*_*t*_, are isolated and do not participate in the infection dynamics. The base infection rate, defined as the probability that a suseptible individual gets infected when she comes in contact with an infected individual, is 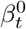. A susceptible individual at time *t* get infected at time *t* + 1 if the susceptibe individual comes in contact with one or more infected individual. The probability that a susceptible individual *i* at time *t* becomes infected at time *t* + 1, denoted by P{*i* ∈ 𝒰_*t*+1_|*i* ∈ 𝒮_*t*_} is obtained by considering first, the probability that a susceptible individual meets *k* undetected individuals, where *k* can take values in 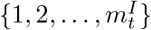 (recall 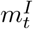 is the internal contact rate); and then, multiplying this probability by the probability of infection and finally, summing over *k*. The number of ways in which a susceptible individual meets *k* infected individuals is given by, 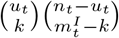, where equals *n*_*t*_ = *s*_*t*_ + *u*_*t*_ + *r*_*t*_. Similarly, the number of ways in which the susceptible individual can meet 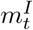 individuals equals 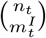. Therefore, the probability that susceptible individual meets *k* infected individual in period *t* is given by

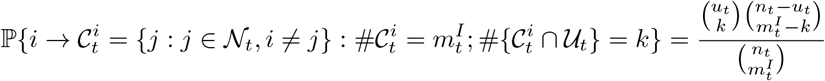

for the individual by *i*. The probability that the susceptible individual gets infected given that the the individual meets *k* infected individuals is given as

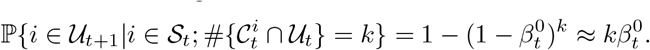

The approximation 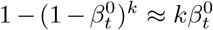 is acceptable since *β*_0_ is usually much smaller than 1, approximately in the range of 0.01−0.05. Therefore, the probability that a susceptible individual meets *k* infected individual and gets infected is given by

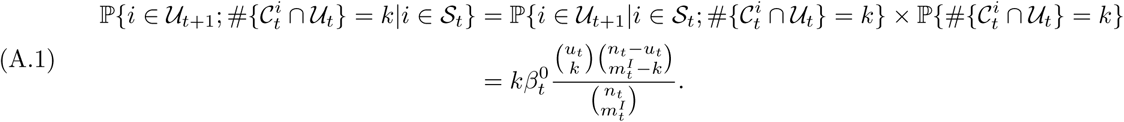

Therefore, the probability a susceptible individual becomes infected at time *t* is

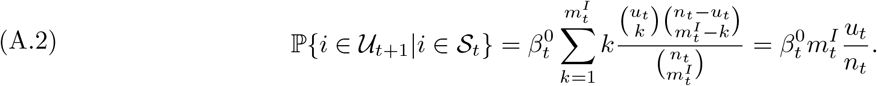

The last expression is obtained by replacing the summation with the expectation of the hypergeometric distribution. The probability of external infection is also similarly obtained as 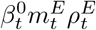, where 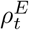 plays the role of *u*_*t*_*/n*_*t*_. Therefore, the total number of new infections in period *t* is given by the product of the number of susceptibles at time *t* with the probability of each susceptible getting infected. Therefore, expected number of new infections in time *t* is given by

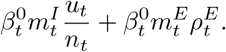

### A.3 Derivation of the probability of infection of an individual among the contact list, 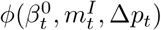

We first derive the probability of infection through within-institutional-transmission (internal) of an individual *i* who is among the contact list and who was not infected in period *t* − 1, i.e., 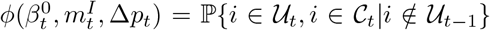. The probability of infection of individuals in the contact list given that the individual was not infected in the previous period is derived by considering that individual *i* can meet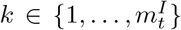 infected individuals, of which at least one individual needs to be among the detected positive cases Δ*p*_*t*_ that is the individual *i* is in the contact list of the detected individuals. Recall that the probability of infection for an individual in the contact list is obtained by adding the probability that the individual was already infected in the previous period, and the probability that the individual is newly infected after the previous period, i.e., 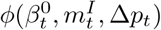. Also, for simplicity, in the following expressions we ignore the notation that the individual *i* was not infected in the previous period. Therefore, we have the following expression:

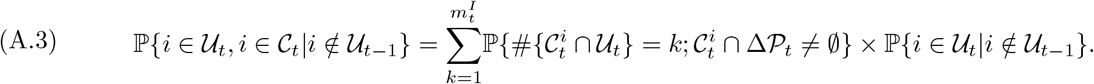

The term P{*i* ∈ 𝒰_*t*_|*i* ∈*/* 𝒰_*t*−1_} is derived as above and equals 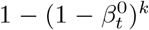. The probability of meeting *k* individuals of which at least one is in Δ*p*_*t*_ is given by

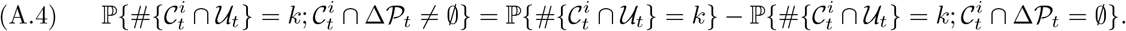

The probability of meeting *k* individuals in a small population is modeled using the Hypergeometric probability. The mobile population *n*_*t*_ = *s*_*t*_ + *u*_*t*_ + *r*_*t*_ at time *t* is divided into two groups, (i) undetected infected group, *u*_*t*_, who can transmit infection, and (ii) the susceptible and recovered group, *s*_*t*_ + *r*_*t*_, who cannot transmit infection further. Therefore, the first probability, 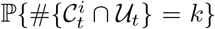, is given by the ratio of the product of the number of ways of choosing *k* from *u*_*t*_ possibilities and choosing the rest 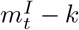 from *s*_*t*_ + *r*_*t*_ group, to the number of ways of choosing the total 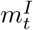 contacts from *n*_*t*_ individuals. Similarly, the second probability, 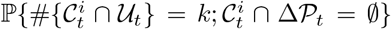, is obtained by the ratio of the product of the number of ways of choosing *k* infected individuals from *u*_*t*_ − Δ*p*_*t*_ individuals (since, we are interested in the probability of detecting contacts with infected individuals who are not in the detected group, Δ*p*_*t*_), the number of ways of choosing 0 individuals from Δ*p*_*t*_ newly detected infected individuals, and choosing the rest of the 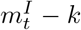 contacts from *s*_*t*_ + *r*_*t*_ individuals, to the number of ways of choosing the total 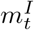 individuals from *n*_*t*_ individuals. Thus, we have

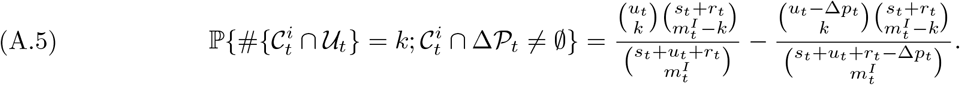

Therefore, combining the above, the function 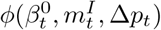 is

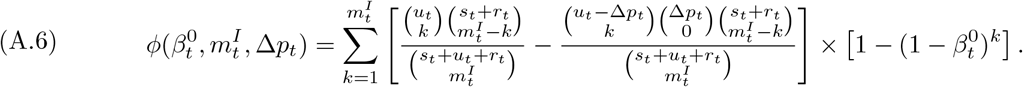

However, the above expression considers only infections through internal transmission. However, individuals in the contact list can also get infected by external sources. Therefore, to extend the function 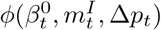 to include infections from outside the organizations, the expression becomes (after considering that 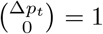)

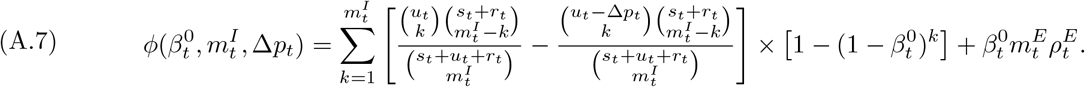

We consider the regime where 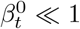 and 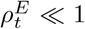. This is acceptable since the infectivities are usually of the order of 5%, and the maximum environmental positivty is below 10%. Therefore, this assumption holds for the case of COVID-19. This assumption leads to the approximation 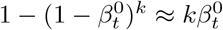, that gives

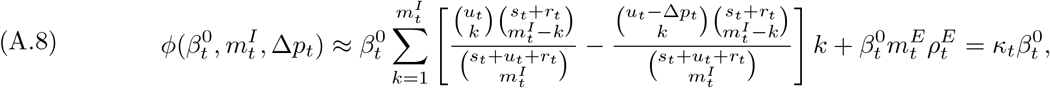

where, we have

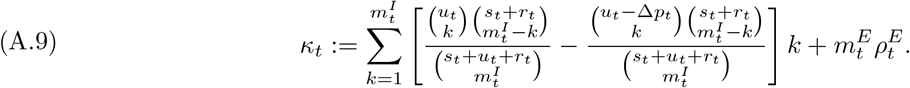

#### A.3.1. Approximating κ_*t*_

From the above derivation, we get

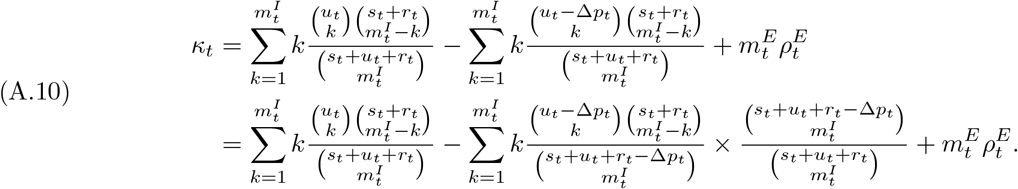

As mentioned earlier, for analytical simplicity, we consider the regime where Δ*p*_*t*_ « *s*_*t*_ + *u*_*t*_ + *r*_*t*_ = *n*_*t*_, where recall that *n*_*t*_ is the size of the mobile part of the population. This is acceptable because in the case of UIUC data, we observe that the maximum detected Δ*p*_*t*_ < 500 in a population of 50K individuals.

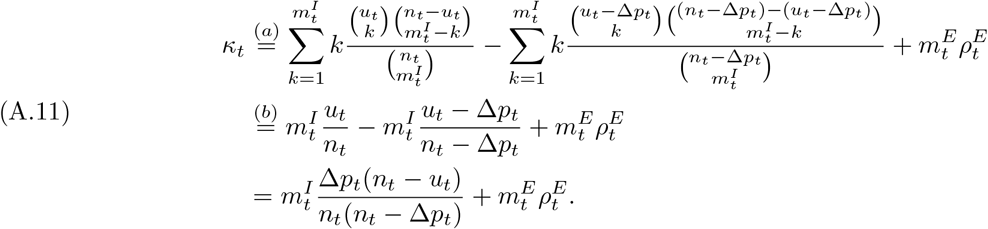

The step in (b) follows from noting that the first summation in (a) is the expectation of a hypergeometric distribution with parameters 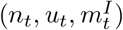, while the second one is the same for a hypergeometric distribution with parameters 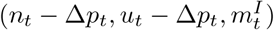.

#### A.3.2. Derivation of the contact traced positive individuals

As indicated in the earlier section, the probability of an individual in the contact list is equal to the prior probability of infection, which is given BY 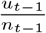, and the probability of getting infected in day *t*, given that the individual was not infected earlier. The probability that an individual in the contact list is infected on day *t* given that the individual was not infected earlier is derived in the previous section as 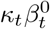, where, the value of *κ*_*t*_, as we have derived earlier in (A.11). The probability that an individual was not infected earlier is given by 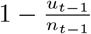. Therefore, the total probability of infection of a contact traced individual is given by

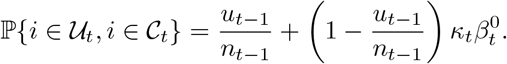

## Appendix B. List of Universities in the Empirical Dataset with Location Information

**Table 8.**
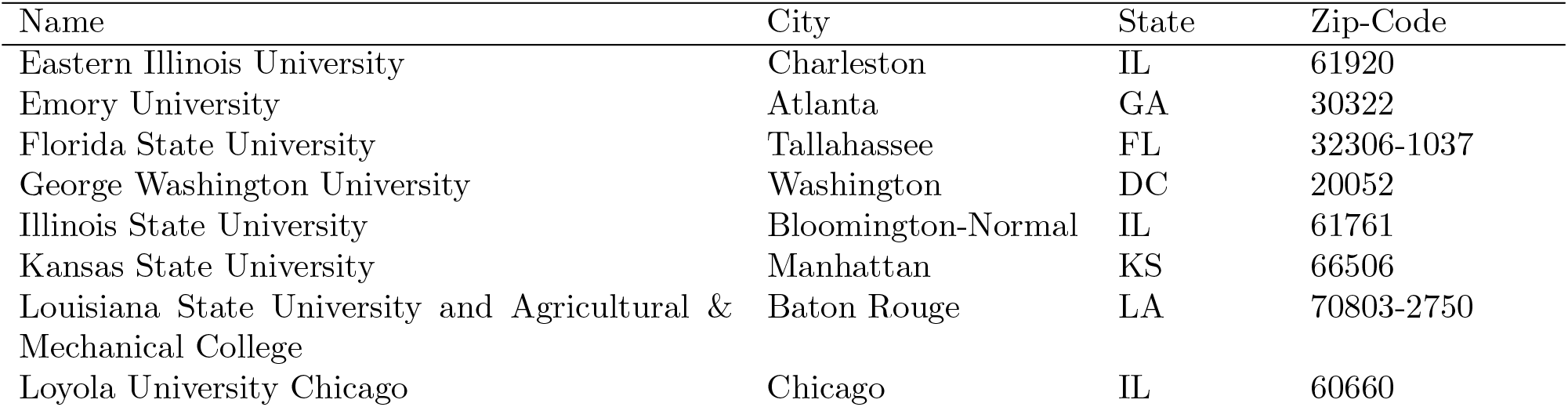
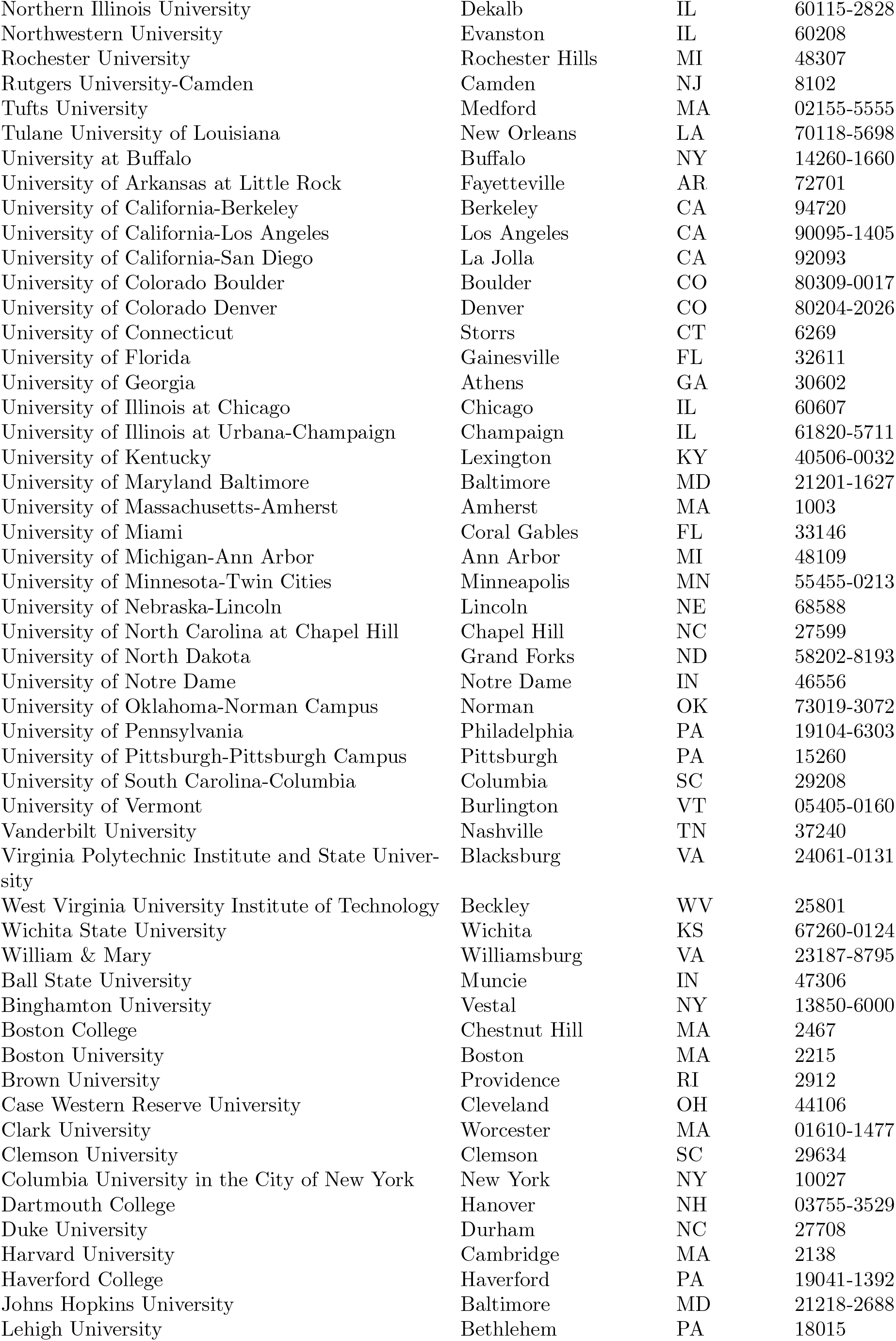
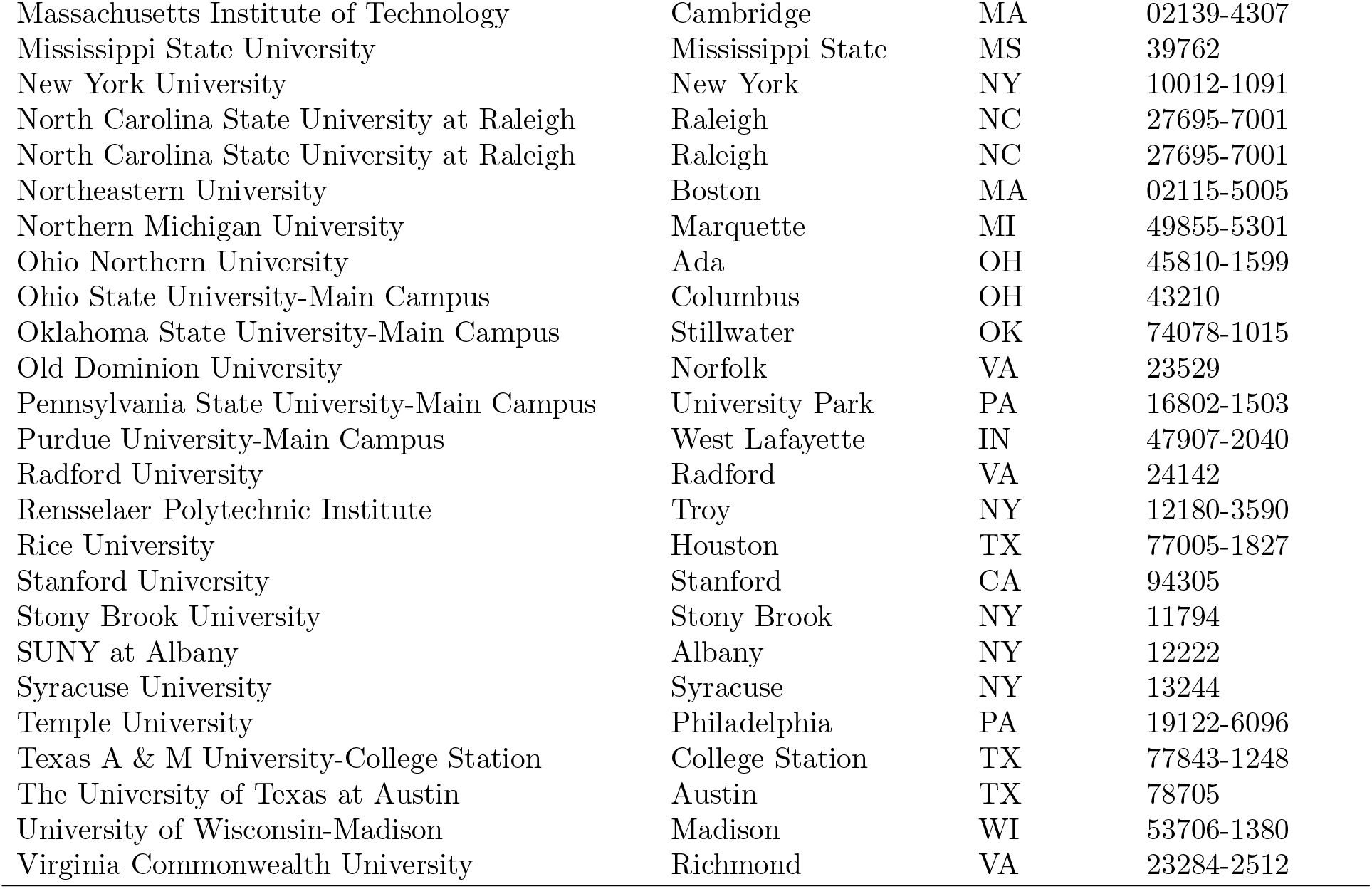
List of Universities with location.

| Name | City | State | Zip-Code |
| --- | --- | --- | --- |
| Eastern Illinois University | Charleston | IL | 61920 |
| Emory University | Atlanta | GA | 30322 |
| Florida State University | Tallahassee | FL | 32306-1037 |
| George Washington University | Washington | DC | 20052 |
| Illinois State University | Bloomington-Normal | IL | 61761 |
| Kansas State University | Manhattan | KS | 66506 |
| Louisiana State University and Agricultural & Mechanical College | Baton Rouge | LA | 70803-2750 |
| Loyola University Chicago | Chicago | IL | 60660 |
| Northern Illinois University | Dekalb | IL | 60115-2828 |
| Northwestern University | Evanston | IL | 60208 |
| Rochester University | Rochester Hills | MI | 48307 |
| Rutgers University-Camden | Camden | NJ | 8102 |
| Tufts University | Medford | MA | 02155-5555 |
| Tulane University of Louisiana | New Orleans | LA | 70118-5698 |
| University at Buffalo | Buffalo | NY | 14260-1660 |
| University of Arkansas at Little Rock | Fayetteville | AR | 72701 |
| University of California-Berkeley | Berkeley | CA | 94720 |
| University of California-Los Angeles | Los Angeles | CA | 90095-1405 |
| University of California-San Diego | La Jolla | CA | 92093 |
| University of Colorado Boulder | Boulder | CO | 80309-0017 |
| University of Colorado Denver | Denver | CO | 80204-2026 |
| University of Connecticut | Storrs | CT | 6269 |
| University of Florida | Gainesville | FL | 32611 |
| University of Georgia | Athens | GA | 30602 |
| University of Illinois at Chicago | Chicago | IL | 60607 |
| University of Illinois at Urbana-Champaign | Champaign | IL | 61820-5711 |
| University of Kentucky | Lexington | KY | 40506-0032 |
| University of Maryland Baltimore | Baltimore | MD | 21201-1627 |
| University of Massachusetts-Amherst | Amherst | MA | 1003 |
| University of Miami | Coral Gables | FL | 33146 |
| University of Michigan-Ann Arbor | Ann Arbor | MI | 48109 |
| University of Minnesota-Twin Cities | Minneapolis | MN | 55455-0213 |
| University of Nebraska-Lincoln | Lincoln | NE | 68588 |
| University of North Carolina at Chapel Hill | Chapel Hill | NC | 27599 |
| University of North Dakota | Grand Forks | ND | 58202-8193 |
| University of Notre Dame | Notre Dame | IN | 46556 |
| University of Oklahoma-Norman Campus | Norman | OK | 73019-3072 |
| University of Pennsylvania | Philadelphia | PA | 19104-6303 |
| University of Pittsburgh-Pittsburgh Campus | Pittsburgh | PA | 15260 |
| University of South Carolina-Columbia | Columbia | SC | 29208 |
| University of Vermont | Burlington | VT | 05405-0160 |
| Vanderbilt University | Nashville | TN | 37240 |
| Virginia Polytechnic Institute and State University | Blacksburg | VA | 24061-0131 |
| West Virginia University Institute of Technology | Beckley | WV | 25801 |
| Wichita State University | Wichita | KS | 67260-0124 |
| William & Mary | Williamsburg | VA | 23187-8795 |
| Ball State University | Muncie | IN | 47306 |
| Binghamton University | Vestal | NY | 13850-6000 |
| Boston College | Chestnut Hill | MA | 2467 |
| Boston University | Boston | MA | 2215 |
| Brown University | Providence | RI | 2912 |
| Case Western Reserve University | Cleveland | OH | 44106 |
| Clark University | Worcester | MA | 01610-1477 |
| Clemson University | Clemson | SC | 29634 |
| Columbia University in the City of New York | New York | NY | 10027 |
| Dartmouth College | Hanover | NH | 03755-3529 |
| Duke University | Durham | NC | 27708 |
| Harvard University | Cambridge | MA | 2138 |
| Haverford College | Haverford | PA | 19041-1392 |
| Johns Hopkins University | Baltimore | MD | 21218-2688 |
| Lehigh University | Bethlehem | PA | 18015 |
| Massachusetts Institute of Technology | Cambridge | MA | 02139-4307 |
| Mississippi State University | Mississippi State | MS | 39762 |
| New York University | New York | NY | 10012-1091 |
| North Carolina State University at Raleigh | Raleigh | NC | 27695-7001 |
| North Carolina State University at Raleigh | Raleigh | NC | 27695-7001 |
| Northeastern University | Boston | MA | 02115-5005 |
| Northern Michigan University | Marquette | MI | 49855-5301 |
| Ohio Northern University | Ada | OH | 45810-1599 |
| Ohio State University-Main Campus | Columbus | OH | 43210 |
| Oklahoma State University-Main Campus | Stillwater | OK | 74078-1015 |
| Old Dominion University | Norfolk | VA | 23529 |
| Pennsylvania State University-Main Campus | University Park | PA | 16802-1503 |
| Purdue University-Main Campus | West Lafayette | IN | 47907-2040 |
| Radford University | Radford | VA | 24142 |
| Rensselaer Polytechnic Institute | Troy | NY | 12180-3590 |
| Rice University | Houston | TX | 77005-1827 |
| Stanford University | Stanford | CA | 94305 |
| Stony Brook University | Stony Brook | NY | 11794 |
| SUNY at Albany | Albany | NY | 12222 |
| Syracuse University | Syracuse | NY | 13244 |
| Temple University | Philadelphia | PA | 19122-6096 |
| Texas A & M University-College Station | College Station | TX | 77843-1248 |
| The University of Texas at Austin | Austin | TX | 78705 |
| University of Wisconsin-Madison | Madison | WI | 53706-1380 |
| Virginia Commonwealth University | Richmond | VA | 23284-2512 |

## Acknowledgements

We acknowledge the support of C3.ai Digital Transformation Institute in funding this research. We also acknowledge the support of the UIUC SHIELD program and program director Ronald Watkins for providing inputs and encouragement for this work.

## Data Repository

The data and codes for this program are available in the following websites:

1. Summary description of the project is available at https://calm-savannah-94927.herokuapp.com,
2. UIUC data is available at https://covid19.illinois.edu/on-campus-covid-19-testing-data-dashboard/,
3. Data for all other 85 universities are available at www.covidedutrends.com,
4. The simulation codes are available at https://github.com/heart-group/institutions-model,
5. A preprint version of the paper is available at https://www.medrxiv.org/content/10.1101/2020.09.04.20188680v4.

## Footnotes

1 To extend our results to organizations outside of educational institutions, one might need different *β*^0^’s for contacts within and outside the organization, depending on the nature of the jobs within that organization.

